# Clinical and neurophysiological determinants of response to contralesional low-frequency repetitive transcranial magnetic stimulation after stroke: A systematic review and meta-analysis

**DOI:** 10.64898/2026.08.20.26360649

**Authors:** Mianxuan Yu, Yiming Zeng, Huanghao Zhou, Jun Lin, Man Hao

## Abstract

**Background:** Low-frequency repetitive transcranial magnetic stimulation (LF-rTMS) over the contralesional primary motor cortex is widely used for post-stroke upper-limb rehabilitation, but treatment response varies substantially. This systematic review and meta-analysis aimed to quantify the efficacy of contralesional LF-rTMS and to examine whether baseline motor impairment severity and corticospinal tract (CST) integrity modify treatment effects.

**Methods:** We searched seven databases from inception to July 2026 for randomized controlled trials of contralesional LF-rTMS (≤1 Hz) versus sham after stroke, with comparable rehabilitation in both arms. The primary outcome was the change in Fugl-Meyer Assessment for the upper extremity (FMA-UE) scores. Random-effects meta-analysis used restricted maximum likelihood estimation with Knapp–Hartung adjustment. Effect modification was examined through meta-regression and biomarker-stratified analyses, and neurophysiological outcomes were also synthesized.

**Results:** Thirty trials (33 comparisons, 1,668 participants) were included. LF-rTMS produced greater FMA-UE improvement than sham (mean difference 4.11 points, 95% CI 2.83–5.39; Hedges g 0.64, 95% CI 0.45–0.84), with substantial heterogeneity. Baseline severity did not significantly modify the effect in continuous meta-regression. However, exploratory within-trial biomarker-stratified analyses suggested larger effects in participants with preserved CST integrity or positive motor-evoked potential (MEP) status. LF-rTMS also shortened MEP latency and central motor conduction time, but these measures could not be validated as surrogate endpoints.

**Conclusions:** Contralesional LF-rTMS provides a statistically significant but modest improvement in post-stroke upper-limb motor recovery. Baseline clinical severity alone may not identify responders, whereas CST integrity is an exploratory, hypothesis-generating candidate biomarker. It requires confirmation in adequately powered biomarker-stratified trials before it can inform clinical decisions.

**Trial Registration:** The study was registered with the International Prospective Register of Systematic Reviews (PROSPERO: CRD420261441561).

## Introduction

Stroke remains a leading cause of long-term disability worldwide [1]. Upper extremity motor impairment affects 35% to 69% of survivors and reduces independence and quality of life [2,3]. Despite advances in rehabilitation, many patients recover upper limb function only partially. Effective adjunctive interventions are therefore needed [4].

Repetitive transcranial magnetic stimulation (rTMS) is a promising neuromodulation technique for post-stroke motor rehabilitation. Its rationale rests on the interhemispheric inhibition (IHI) model. Stroke disrupts the balance of reciprocal inhibition between the two hemispheres. The contralesional (unaffected) hemisphere then exerts excessive inhibition on the ipsilesional (affected) hemisphere, which impedes motor recovery [5]. Low-frequency rTMS (LF-rTMS, ≤1 Hz) over the contralesional primary motor cortex (M1) is thought to reduce this abnormal hyperexcitability. This may rebalance interhemispheric inhibition and support recovery of the affected hemisphere [6]. Mechanistic studies show that inhibitory stimulation induces pre- and postsynaptic changes in GABAergic neurotransmission. These findings provide a synaptic basis for contralesional inhibitory protocols [7,8].

A growing number of randomized controlled trials (RCTs) and meta-analyses have examined LF-rTMS for post-stroke upper limb recovery. Earlier syntheses generally support a beneficial effect. A 2025 network meta-analysis [9] ranked LF-rTMS second among seven NIBS techniques for ADL (Barthel Index), behind anodal tDCS. A recent meta-analysis concluded that moderate- to high-quality evidence supports its efficacy for motor function in early stroke [10]. However, heterogeneity across studies is substantial, and several key questions remain unresolved.

First, the role of baseline motor impairment severity as a predictor of treatment response is controversial [11,12]. Some studies suggest that patients with milder impairment benefit more [11]. Others report efficacy in severe hemiparesis [13]. This inconsistency may partly reflect differences in corticospinal tract (CST) integrity. CST integrity is assessed by motor-evoked potential (MEP) status or diffusion tensor imaging (DTI). It is increasingly recognized as a key determinant of both spontaneous recovery and treatment responsiveness [14,15]. Expert consensus has identified MEP status and DTI-derived measures as promising biomarkers. They may characterize corticospinal integrity and predict responsiveness to neurorehabilitation after stroke [16]. Nevertheless, whether CST integrity modifies the effect of LF-rTMS has not been systematically examined.

Second, LF-rTMS modulates neurophysiological measures such as MEP amplitude, latency, and central motor conduction time (CMCT) [17]. Whether these changes are valid surrogate endpoints for clinically meaningful motor recovery remains unknown. Establishing surrogacy would have important implications for trial design and mechanistic inference.

Third, the proportional recovery rule has not been applied at the trial level. This rule posits that stroke patients recover approximately 70% of their maximum potential motor function within 3–6 months, contingent on CST integrity [18,19]. It has been validated in observational cohorts. However, it has not been used to quantify the proportion of recovery potential realized by an active intervention. Such an approach could offer a novel perspective on efficacy that accounts for baseline impairment severity.

Given these unresolved issues, we conducted a systematic review and meta-analysis with five objectives. The first was to quantify the efficacy of contralesional LF-rTMS (≤1 Hz) on upper limb motor recovery. Efficacy was measured by the Fugl-Meyer Assessment for the upper extremity (FMA-UE) and benchmarked against the minimal clinically important difference. The second was to examine whether baseline motor impairment severity modifies the treatment effect. We used continuous meta-regression and prespecified severity bands for this analysis. The third was to explore whether CST integrity modifies the effect, by pooling within-trial stratified data. The fourth was to evaluate whether neurophysiological changes are associated with functional recovery.

The fifth was to apply a proportional-recovery framework, quantifying the proportion of residual recovery potential realized by LF-rTMS.

We restricted the analysis to sham-controlled RCTs of LF-rTMS (≤1 Hz) over the contralesional M1. This isolates the net effect of the intervention. Studies in which the independent effect of LF-rTMS could not be determined were excluded.

## Materials and methods

This review is registered with the International Prospective Register of Systematic Reviews (PROSPERO, CRD420261441561). It follows the Cochrane Handbook for Systematic Reviews of Interventions and is reported in line with the Preferred Reporting Items for Systematic Reviews and Meta-Analyses (PRISMA) 2020 statement [20].This review did not require ethical approval as it synthesized data from previously published studies.

### Literature search strategy

We searched seven Chinese and English databases: PubMed, Embase, Web of Science, the Cochrane Library, China National Knowledge Infrastructure (CNKI), Wanfang, and VIP. The search covered the period from inception to July 6, 2026. Search terms combined Medical Subject Headings (MeSH) and free-text words. Key terms included “stroke,” “upper extremity,” “transcranial magnetic stimulation,” and “randomized controlled trial.” A supplementary search on July 24, 2026, used expert consultation to identify additional eligible studies. This search yielded one additional study, which was included after full-text screening [13]. Only articles published in English or Chinese were eligible. The detailed search strategy is presented in Table S1.

### Eligibility criteria

#### Inclusion criteria

We included randomized controlled trials (RCTs) that met the following criteria: Participants (P). Adults (≥18 years) with clinically and radiologically confirmed ischemic or hemorrhagic stroke were eligible. They had to present with upper limb motor impairment. Studies enrolling patients in the subacute (>14 days to 6 months) or chronic (>6 months) phase after stroke were included.

Interventions (I). Studies evaluating low-frequency repetitive transcranial magnetic stimulation (LF-rTMS, ≤1 Hz) applied over the contralesional primary motor cortex (M1).

Comparison (C). Sham rTMS with otherwise comparable rehabilitation. Studies using stratified randomization were eligible if each stratum could be treated as an independent comparison.

Outcomes (O). The primary outcome was the change score of the Fugl-Meyer Assessment for the upper extremity (FMA-UE). Studies not reporting FMA-UE were ineligible for the quantitative synthesis. Secondary outcomes comprised neurophysiological measures.

Study design (SD). Randomized, double- or single-blind, sham-controlled trials published as full-text articles in English or Chinese.

#### Exclusion criteria

We excluded studies that: (1) involved bilateral stroke or isolated brainstem or cerebellar lesions; (2) used stimulation protocols other than LF-rTMS over contralesional M1; (3) were crossover trials without sufficient washout periods; or (4) lacked a sham rTMS control group. Studies not reporting FMA-UE were excluded from the quantitative synthesis. Non-English or non-Chinese publications were also excluded, as were conference abstracts without a full-text article.

### Study selection

All retrieved records were imported into NoteExpress for management and duplicate removal. Two reviewers (M.Y. and J.L.) independently screened titles and abstracts against the predefined eligibility criteria. Records were excluded at this stage if they were irrelevant to the topic, unrelated to stroke or rTMS, lacked a control group, or did not report relevant outcomes. When eligibility was unclear from the title and abstract, the full text was retrieved and assessed.

The same two reviewers independently evaluated the full texts and recorded the reasons for exclusion. Disagreements were resolved through joint appraisal of the full text and consensus.

### Outcome measures of interest

The primary outcome was the change in upper limb motor function, measured by the Fugl-Meyer Assessment for the upper extremity (FMA-UE) [21]. The FMA-UE is a validated scale for post-stroke upper limb motor impairment. It comprises 33 items covering motor function, reflex activity, coordination, and movement speed. Scores range from 0 to 66, and higher scores indicate better motor performance.

For effect-modification analyses, baseline FMA-UE severity was treated as a potential effect modifier. Study-level mean baseline FMA-UE scores were analyzed as a continuous variable. They were also categorized into predefined severity bands (<20, 20– 30, 30–40, and ≥40) [22].

Secondary outcomes were neurophysiological measures of corticospinal excitability and conduction. These included motor-evoked potential (MEP) amplitude, MEP latency, and central motor conduction time (CMCT). They were extracted when available to explore the neurophysiological effects of LF-rTMS.

Corticospinal tract (CST) integrity, assessed by MEP status or diffusion-based neuroimaging measures, was analyzed as an exploratory effect modifier. This analysis aimed to generate hypotheses about potential predictors of treatment response.

### Data extraction

One reviewer (M.Y.) extracted the data, and the second reviewer (J.L.) independently verified them. Extracted items covered study characteristics (first author, publication year, country, language, design, registration, and ethics approval). They also covered participant characteristics (sample size, stroke type, age, sex, time since stroke, and baseline motor impairment). Intervention details included rTMS frequency, intensity, pulses per session, number of sessions, stimulation site, coil type, navigation method, and concurrent rehabilitation. Control conditions (sham method or conventional rehabilitation) were recorded. Outcome measures (FMA-UE, neurophysiological outcomes, and follow-up data) and randomization and blinding procedures were also recorded.

### Risk of bias assessment

Two reviewers (M.Y. and J.L.) independently assessed the risk of bias of the included trials, using the Cochrane Risk of Bias 2 (RoB 2) tool [23]. The tool evaluates five domains: bias arising from the randomization process; bias due to deviations from intended interventions; bias due to missing outcome data; bias in measurement of the outcome; and bias in selection of the reported result. Each domain was judged as low risk of bias, some concerns, or high risk of bias. Disagreements were resolved through discussion or consultation with a third reviewer (H.Z.).

The certainty of evidence for each outcome was evaluated with the Grading of Recommendations Assessment, Development and Evaluation (GRADE) approach [24]. Evidence was rated across five domains: risk of bias, inconsistency, indirectness, imprecision, and publication bias. Overall certainty was categorized as high, moderate, low, or very low.

### Data synthesis and statistical analysis

All analyses were performed using Python 3.11 (NumPy, pandas, and SciPy). Statistical heterogeneity was assessed with the I² statistic and τ². I² ≥ 50% was taken to indicate substantial heterogeneity. Given the anticipated clinical and methodological diversity, a restricted maximum-likelihood (REML) random-effects model was used as the primary approach. The Knapp–Hartung small-sample adjustment was applied [25]. DerSimonian– Laird and Paule–Mandel estimators were computed as sensitivity analyses. Ninety-five percent prediction intervals were calculated for syntheses with ≥10 comparisons.

When change scores between baseline and end of treatment were reported, they were used directly. Otherwise, change scores were calculated from baseline and post-treatment means. The standard deviation (SD) of the change was imputed with a pre–post correlation of r = 0.5, following the Cochrane Handbook (sensitivity analyses: r = 0.2 and r = 0.8). Of the 33 comparisons, 25 required imputation of the change-score SD. One further comparison used a conservatively imputed value for an anomalous reported SD (implied r > 0.95). Four studies (8 arms) reported complete baseline, post-treatment, and change SDs. Their empirical pre–post correlations had a median of 0.91 (range 0.74– 0.99). The value r = 0.5 is therefore conservative, and the sensitivity range (r = 0.2/0.8) brackets the plausible values. Where a reported change-score SD implied an implausible correlation (r > 0.95), conservative imputed values were used in the primary analysis. The reported values were used in sensitivity analysis. Data reported as standard errors or 95% confidence intervals were converted to SDs with standard formulae. Studies reporting only medians and interquartile ranges were not synthesized quantitatively; they were summarized narratively. For multi-arm trials, eligible low-frequency arms were combined into a single comparison. Sample sizes were summed, means were pooled with arm-size weighting, and SDs were combined with the standard formula for merging groups. This formula incorporates within-arm variances and squared deviations of arm means from the pooled mean. Strata of stratified-randomization trials were entered as independent comparisons.

Standardized mean differences (SMD, Hedges g) and mean differences (MD, FMA-UE points) were pooled with 95% CIs. Effect modification by baseline severity was tested with random-effects meta-regression, using linear and quadratic terms. Prespecified subgroup analyses covered country, stroke phase, and MEP enrichment. For the proportional-recovery analysis, the between-arm difference in FMA-UE change was divided by the distance to the scale ceiling, calculated as (66 − baseline FMA-UE), following the proportional recovery rule [18]. This metric assumes that recovery potential scales with baseline impairment and is most applicable to subacute populations. It was therefore analysed as a secondary, descriptive measure.

All included trials measured the primary outcome on the same scale (FMA-UE, 0–66 points). The mean difference (MD, in FMA-UE points) was therefore used as the primary effect measure to preserve clinical interpretability. It was benchmarked against the minimal clinically important difference (≈5–6 points) [26]. The standardized mean difference (SMD, Hedges g) was calculated for analyses requiring scale independence, such as meta-regression and neurophysiological syntheses. Effect sizes and 95% CIs are presented on forest plots. A two-sided P < 0.05 was considered statistically significant.

The prespecified severity bands were <20, 20–30, 30–40, and ≥40 FMA-UE points [22]. Sensitivity analyses comprised exclusion of high risk-of-bias studies and of the largest-weight trial. They also comprised exclusion of studies with data-quality flags, dual analysis of the anomalous-SD study, alternative imputation correlations, and re-computation under the MD metric.

Publication bias was assessed by visual inspection of funnel-plot symmetry. Formal assessment used Egger’s regression [27] and Peters’ [28] tests when ≥10 comparisons were available. Where Egger’s test was significant or borderline (P < .10), trim-and-fill [29] and PET-PEESE [30] adjustments were applied. The certainty of evidence for each outcome was rated using the GRADE approach.

## Results

### Study selection

A total of 4,181 records were identified from the seven electronic databases. One additional record was identified through expert consultation. Automation tools removed 1,463 duplicate records and conference abstracts. The remaining 2,719 records were screened by title and abstract. Of these, 2,573 records were excluded according to the prespecified exclusion codes (grouped by PICOS).

Consequently, 146 reports underwent full-text review. Eight were excluded as duplicate publications of the same cohort. A further 107 were excluded for PICOS-based reasons. The main reasons were outcome or data not meeting requirements or not extractable, including reporting only medians with interquartile ranges. Other reasons were comparator structure not fitting the criteria, including questionable sham procedures (e.g., 20% output intensity as sham). Further reasons were inadequate study design, including crossover trials and acute-phase populations. Intervention parameters not fitting the criteria and registered protocols without outcome data also led to exclusion. In total, 31 studies were eligible. Thirty studies (33 comparisons) entered the primary quantitative synthesis. One study [31] reported only six-month change scores and was synthesized qualitatively. The selection process is shown in Fig 1.

**Fig 1.**
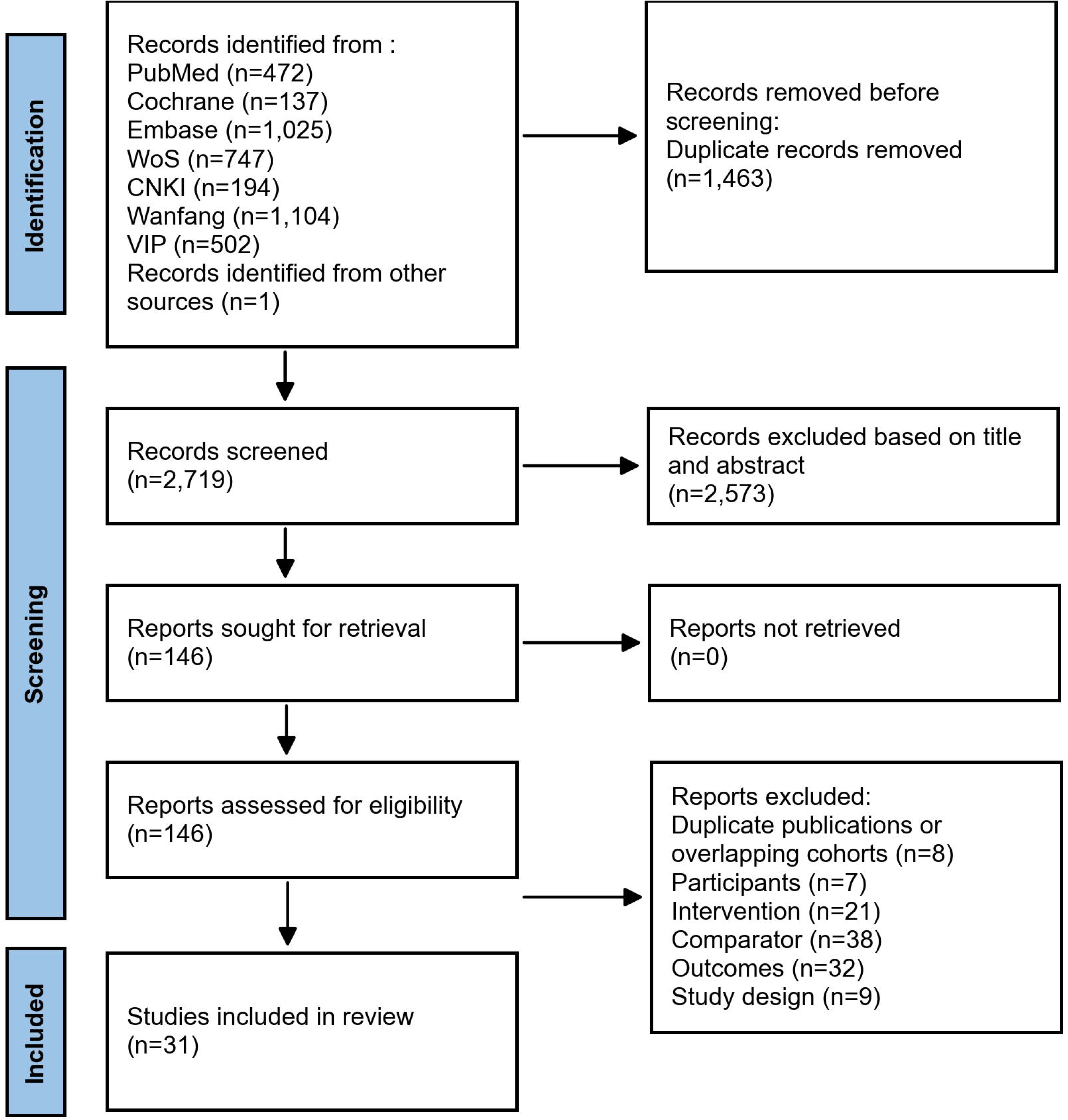
PRISMA flow diagram.

### Characteristics of included studies

The characteristics of the 31 included studies are summarized in Table 1. Thirty RCTs (33 comparisons; 1,668 participants) entered the primary quantitative synthesis. One trial reporting only six-month change scores ([31]; n = 58) was synthesized qualitatively.

**Table 1.**
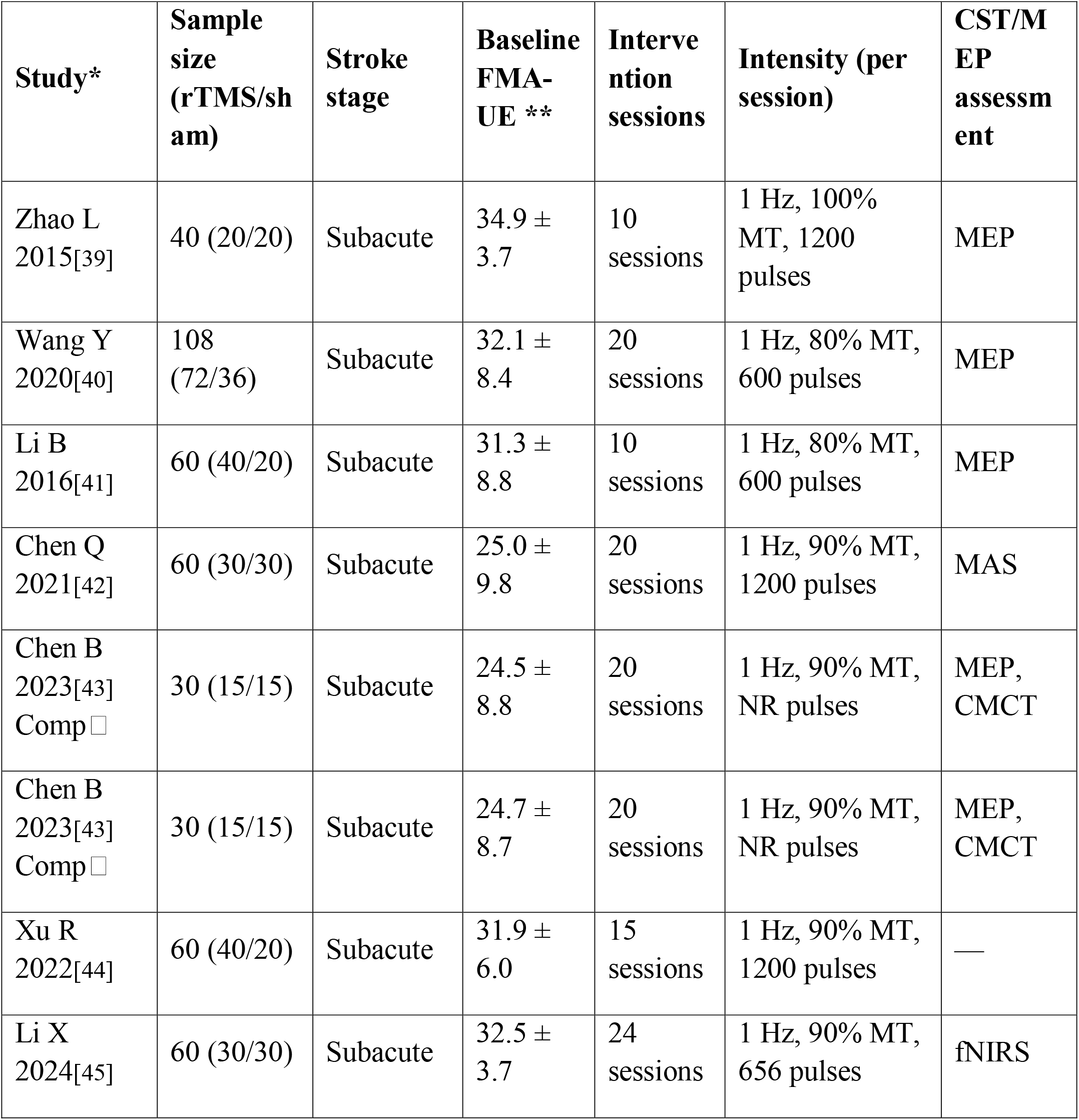
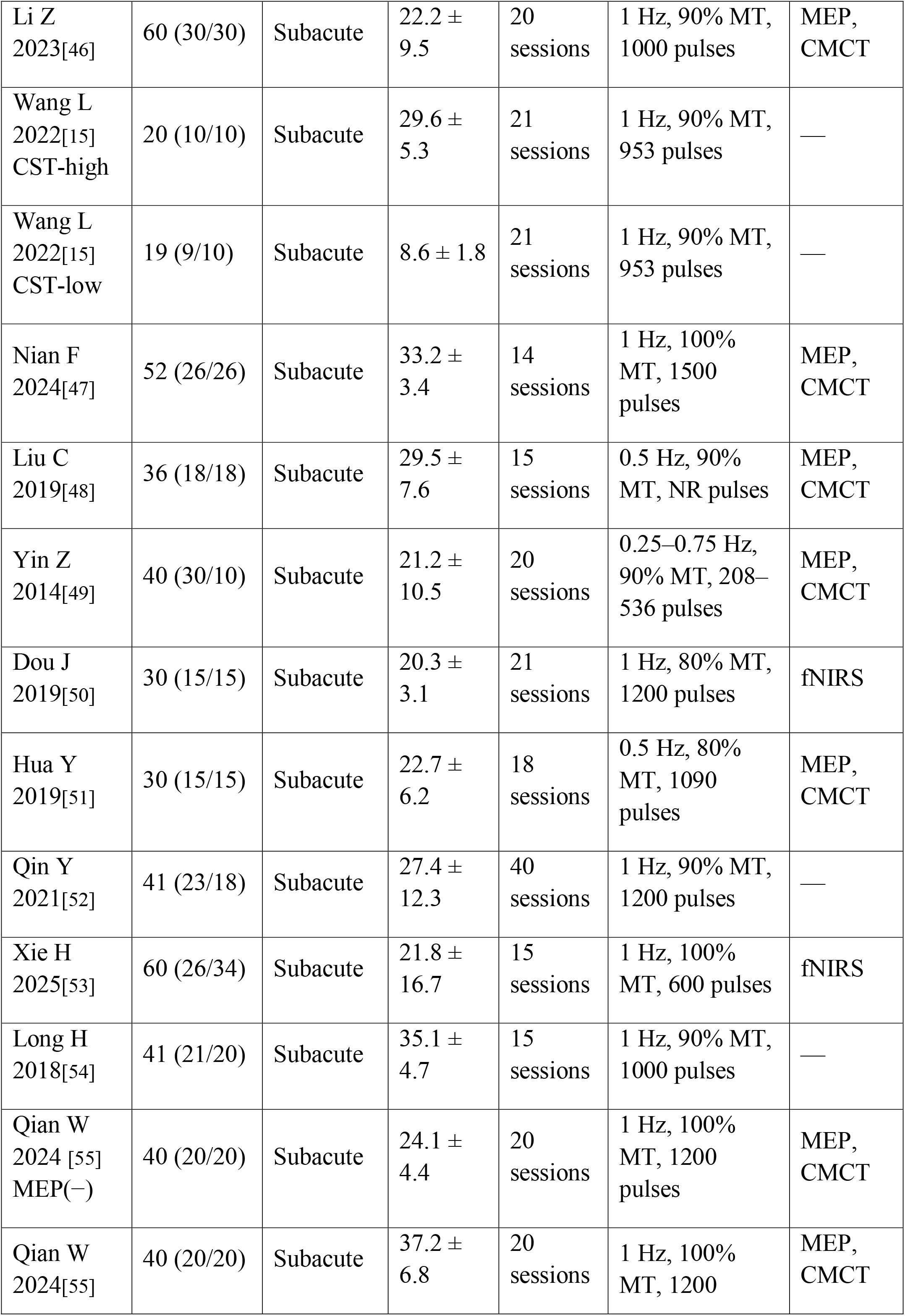
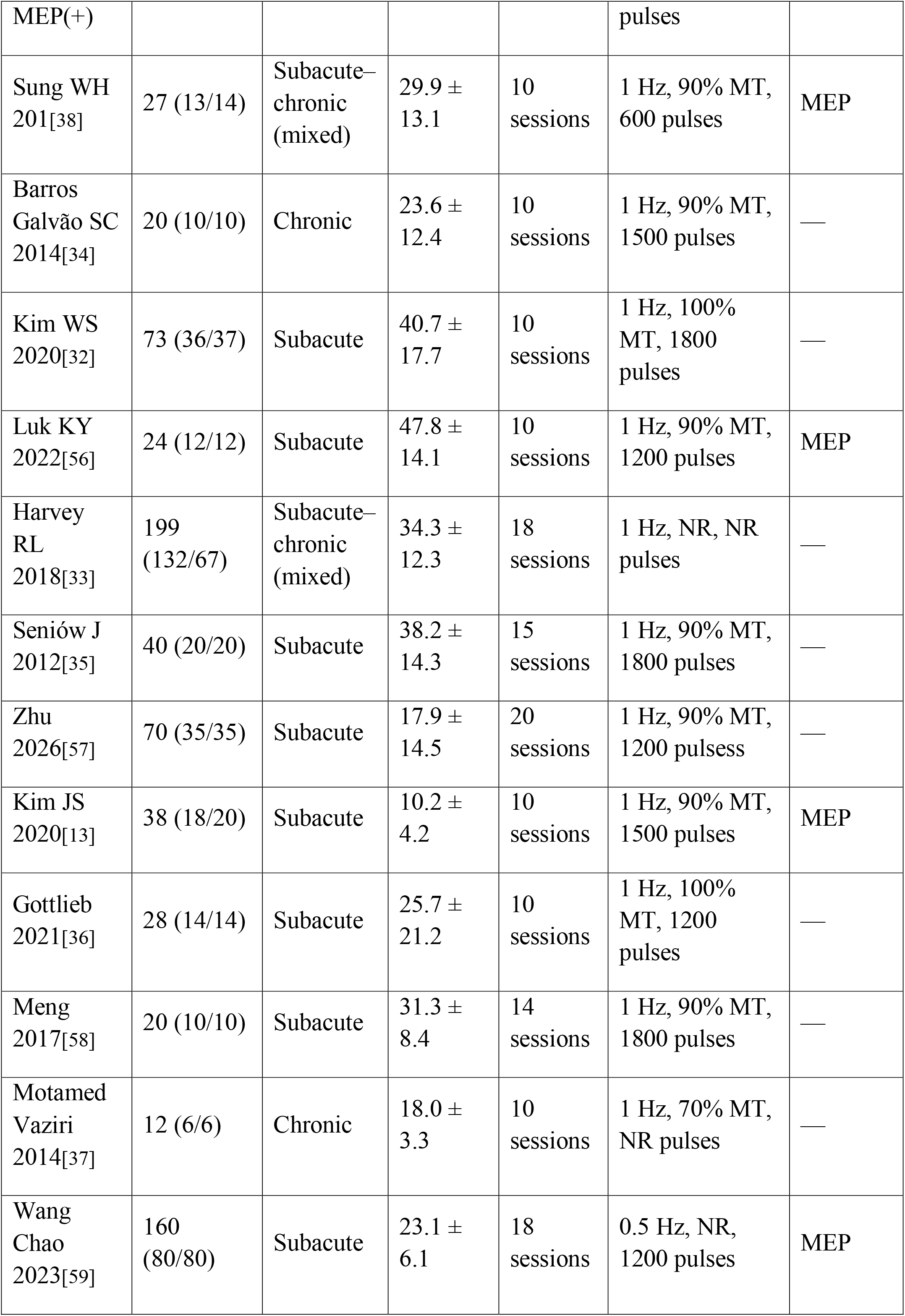
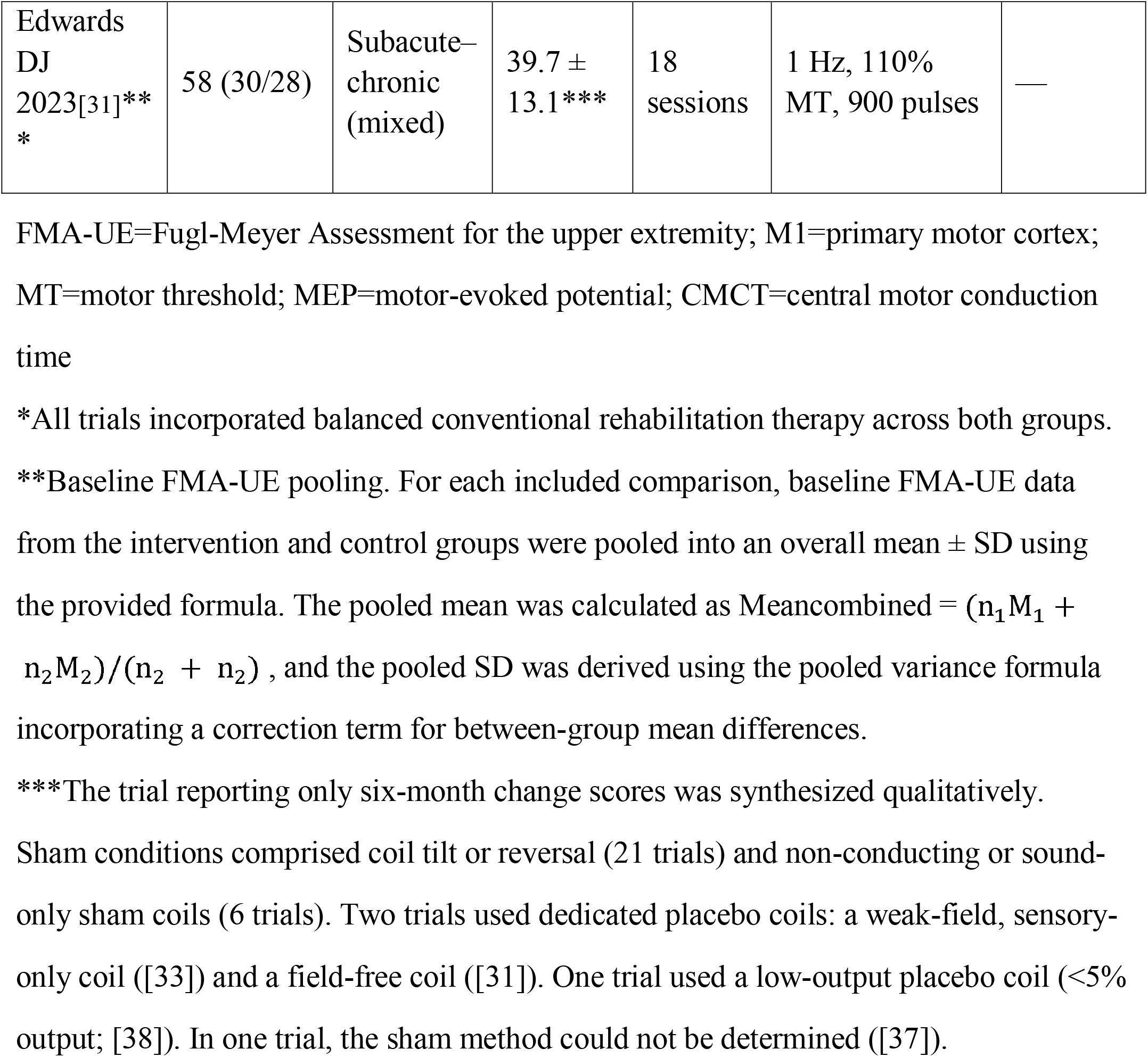
Study characteristics.

Sample sizes ranged from 12 to 199. Most trials were conducted in China (23, including Taiwan and Hong Kong). The remainder came from Korea (2) [13,32], the United States (2) [31,33], Brazil [34], Poland [35], Germany [36], and Iran [37] (1 each). Twenty-six trials enrolled subacute patients, two enrolled chronic patients, and three included mixed subacute-to-chronic populations. Baseline FMA-UE scores ranged from 10.2 to 47.8.

Twelve trials recruited patients with moderate impairment, 12 with moderate-to-severe impairment, 3 with severe impairment, and 2 with mild impairment. Two trials stratified participants by corticospinal tract integrity or motor-evoked potential (MEP) status. All trials applied LF-rTMS over the contralesional primary motor cortex (1 Hz in 27 trials, 0.5 Hz in 3, and 0.25 Hz in 1). Treatment comprised 10–40 sessions (median 15), superimposed on balanced conventional rehabilitation in both arms.

FMA-UE was the primary outcome in all trials; secondary outcomes included neurophysiological measures (MEP, CMCT). Compared with earlier syntheses, this review includes trials published up to July 2026, among them two biomarker-stratified RCTs. To our knowledge, it is the first to prespecify baseline severity as an effect modifier.

### Risk of bias

Among the 31 included studies, two had an overall low risk of bias [56,57]. Both reported adequate randomization, allocation concealment, blinded outcome assessment, and prospective registration. Twenty-three studies raised some concerns. Six studies had a high overall risk of bias [37,41,45,49,50,52] (Fig 2B).

**Fig 2.**
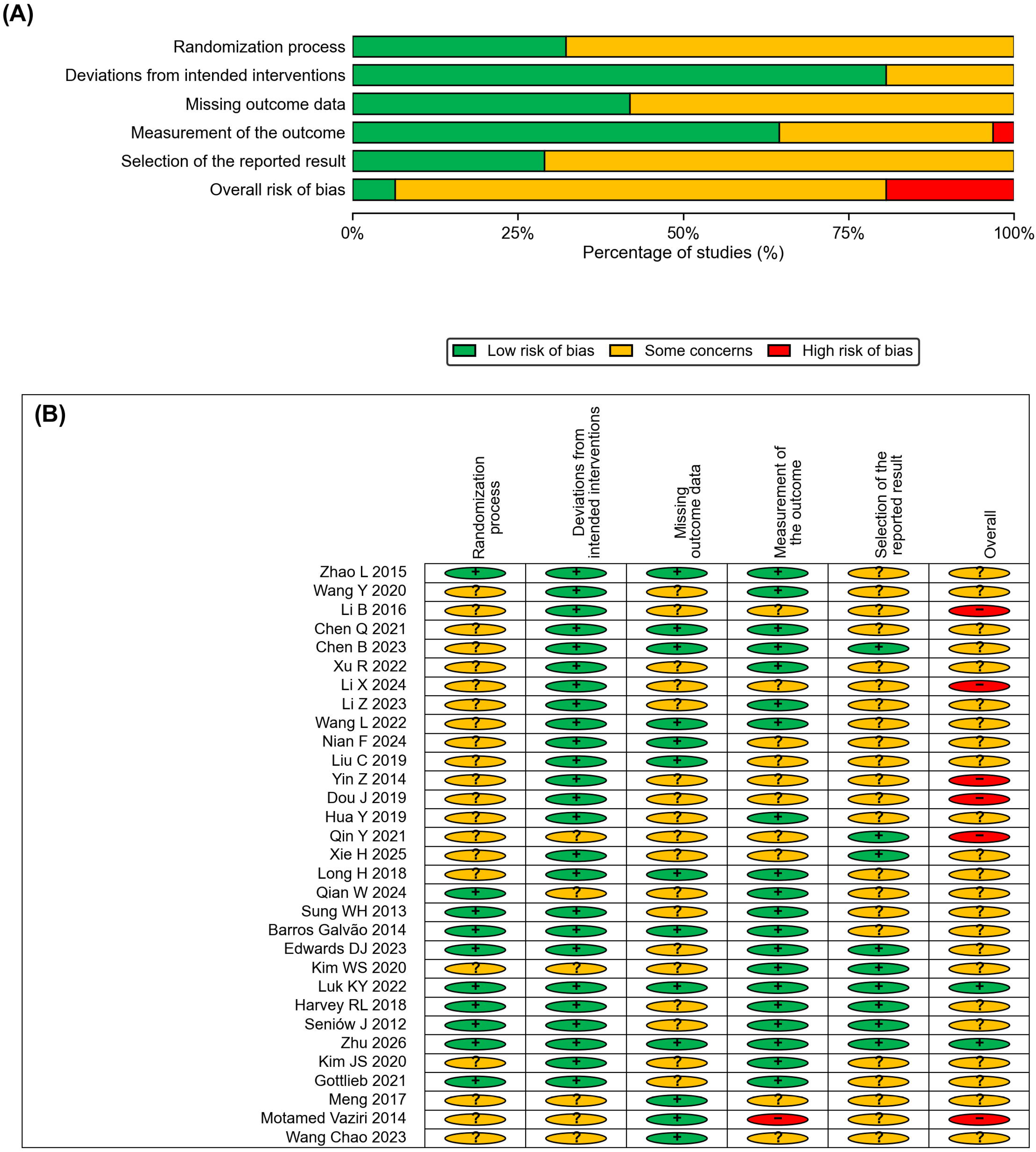
RoB 2 summary.

At the domain level (Fig 2A), concerns were most frequent for selection of the reported result and for missing outcome data. Only nine studies were at low risk for selective reporting, mainly because prospective registration or prespecified analysis plans were rarely reported. Only 13 studies were at low risk for missing outcome data, mainly because attrition, its handling, and intention-to-treat analyses were incompletely documented. Random sequence generation and allocation concealment (D1) were adequate in 10 studies. Deviations from intended interventions (D2) showed comparatively favorable ratings, with 25 studies at low risk. Outcome measurement (D4) was at low risk in 20 studies. One study was at high risk because the outcome assessor was not blinded [37]. Ten further studies did not report whether assessors were blinded. Domain-level judgments are detailed in Table S2.

Overall, the certainty of evidence was limited mainly by methodological concerns. These included incomplete reporting of allocation concealment, attrition handling, and trial registration. Statistical heterogeneity, documented below, also contributed. Outcome-specific GRADE ratings are reported with the corresponding syntheses (Table S3).

### Overall effect of LF-rTMS on upper limb motor recovery

Across 33 comparisons from 30 randomized controlled trials (1,668 participants), contralesional LF-rTMS improved upper-limb motor function more than sham stimulation. Both groups received otherwise comparable rehabilitation (Fig 3).

Fig 3. Forest plot of the effect of contralesional LF-rTMS on FMA-UE change scores. Independent comparisons from the same trial are distinguished by a/b suffixes (e.g., Wang L 2022a/b), as detailed in the figure footnote and Table S1.

In the primary analysis (REML random-effects model with Knapp–Hartung adjustment), the pooled mean difference in FMA-UE change scores was 4.11 points (95% CI 2.83 to 5.39; P < 0.001). The corresponding standardized mean difference (Hedges g) was 0.64 (95% CI 0.45 to 0.84; P < 0.001). Heterogeneity was substantial for both metrics (MD: I² = 81%, τ² = 9.55, Cochran Q = 171.1, df = 32, P < 0.001; SMD: I² = 72%, τ² = 0.20). The 95% prediction interval for the mean difference ranged from −2.32 to 10.54 points. Thus, the average effect clearly favors active stimulation. However, the effect expected in an individual future setting ranges from negligible to well above the clinically important threshold.

The pooled point estimate (4.11 points) fell just below the lower bound of the minimal clinically important difference (approximately 5–6 points) [26]. The upper confidence limit (5.39 points) entered the clinically important range. Twelve of the 33 comparisons (36%) showed between-group differences of at least 5 points. These comparisons were distributed across severity strata rather than concentrated in any single patient subgroup. The single trial synthesized qualitatively [31] reported only six-month change scores. It found no between-group difference at six months (5.17 ± 8.24 versus 5.00 ± 7.29 points). The trial had been terminated early for futility under a prespecified group-sequential rule. Its result was designated inconclusive by the investigators. Possibly, the sham coil produced a widespread weak electric field that affected the cortex.

According to GRADE, the certainty of evidence for the primary outcome was low. The evidence was downgraded once for risk of bias (only 2 of 31 studies at overall low risk). It was downgraded once more for unexplained inconsistency. No downgrade was applied for indirectness, imprecision, or publication bias. The direction of the average effect was consistent across all sensitivity analyses (Table S4).

### Baseline motor impairment severity as a clinical modifier of LF-rTMS response

Baseline motor impairment was examined as a potential clinical modifier because patients with different residual motor capacities may respond differently to neuromodulatory interventions.

#### Continuous meta-regression

Random-effects meta-regression (REML with Knapp–Hartung adjustment) across the 33 comparisons found no linear association between study-level mean baseline FMA-UE and treatment effect (Fig 4). This held on the standardized scale (β = 0.007 per FMA-UE point, 95% CI −0.017 to 0.032; P = 0.544). It also held on the raw scale (β = 0.015 FMA-UE points gained per baseline point, 95% CI −0.140 to 0.169; P = 0.848). Adding a quadratic term did not improve the model (β□ = −0.0002, P = 0.860 for the SMD metric; curvature test on the MD metric, P = 0.170). Thus, there was no support for U-shaped or inverted-U-shaped dose–response patterns. Results were unchanged when the analysis was restricted to the 29 comparisons enrolling exclusively subacute patients (linear term P = 0.474). Rank-based analyses were likewise null (Kendall τ = 0.03, P = 0.79; Spearman ρ = 0.07, P = 0.71). Study-level baseline severity, treated as a continuum, therefore did not detectably modify the treatment effect.

**Fig 4.**
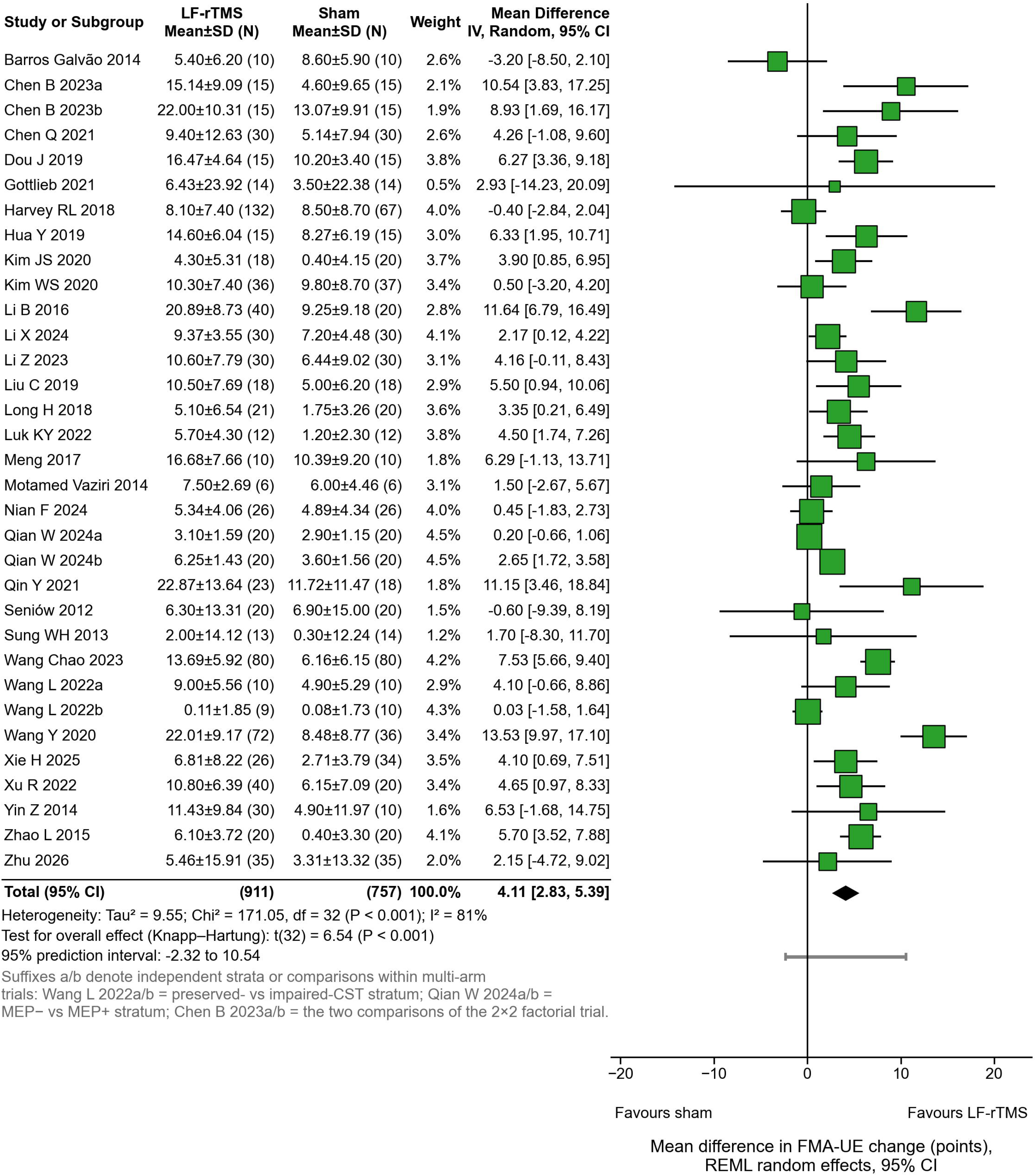
Severity meta-regression and subgroup analyses.

#### Stratified analysis by baseline FMA-UE severity

Stratification into the prespecified severity bands yielded the estimates in Table 2. The pooled effect was smallest in the severe band (baseline FMA-UE < 20; k = 4; SMD 0.33, 95% CI −0.20 to 0.86; I² = 6%). Its confidence interval was wide and compatible with effects from none to moderate. Effects were comparable across the remaining bands. For the 20–30 band (k = 16), the SMD was 0.65 (95% CI 0.41 to 0.89; I² = 56%). For the 30–40 band (k = 11), it was 0.77 (95% CI 0.33 to 1.21; I² = 85%). For the ≥40 band (k = 2), it was 0.60 (95% CI −0.57 to 1.77; I² = 83%). A direct contrast of the severe band against all remaining comparisons was not significant (0.33 versus 0.68; z = −1.79, P = 0.074). Any apparent severity gradient is therefore hypothesis-generating rather than confirmatory. It is also partly confounded by corticospinal tract integrity. Two of the four severe-band comparisons came from biomarker-defined strata with disrupted corticospinal transmission.

**Table 2.**
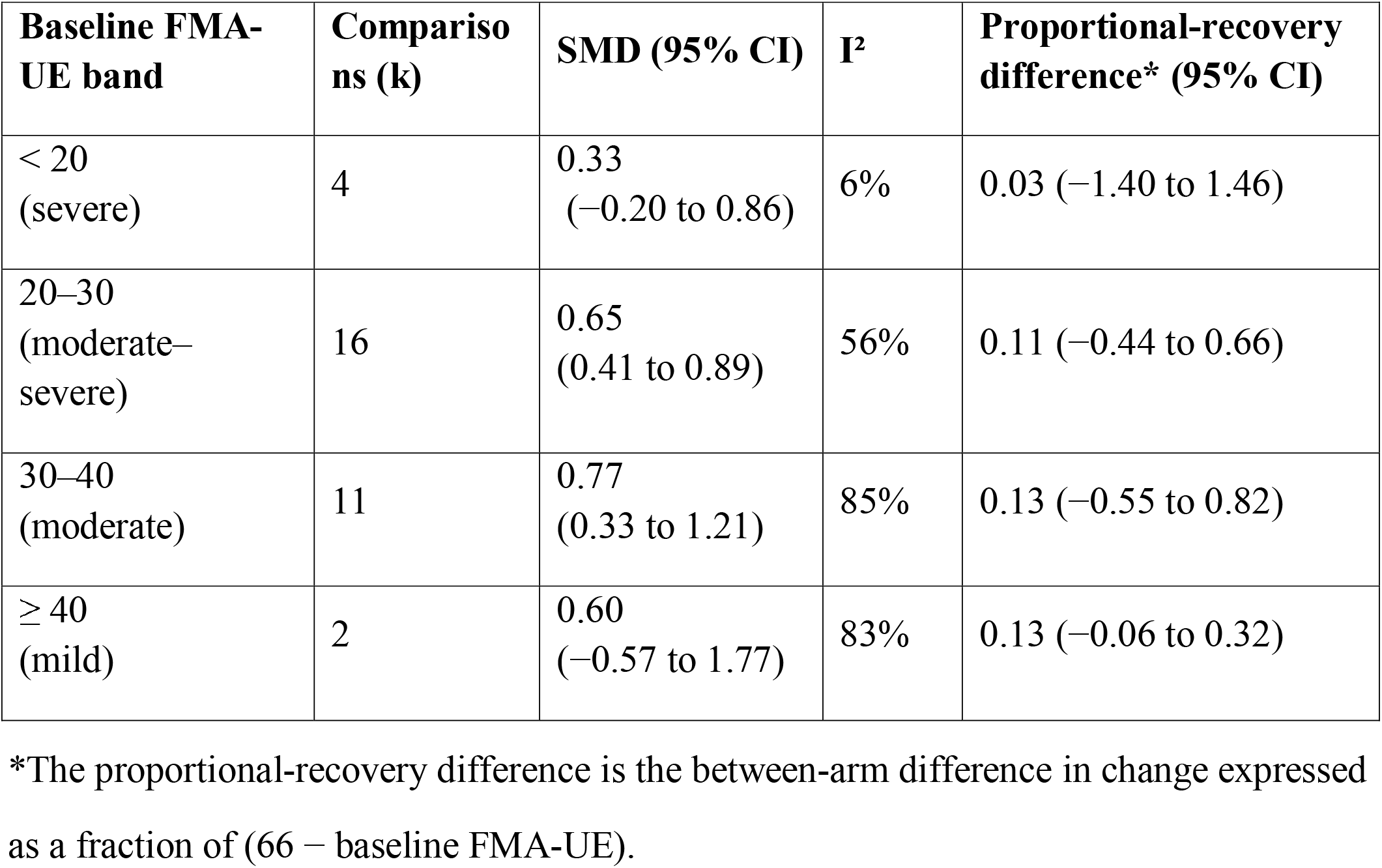
Stratified random-effects meta-analysis of the rTMS effect on FMA-UE change by baseline motor severity (REML with Knapp–Hartung adjustment).

| Baseline FMA-UE band | Comparisons (k) | SMD (95% CI) | I<sup>2</sup> | Proportional-recovery difference* (95% CI) |
| --- | --- | --- | --- | --- |
| < 20<br>(severe) | 4 | 0.33<br>(-0.20 to 0.86) | 6% | 0.03 (-1.40 to 1.46) |
| 20–30<br>(moderate–severe) | 16 | 0.65<br>(0.41 to 0.89) | 56% | 0.11 (-0.44 to 0.66) |
| 30–40<br>(moderate) | 11 | 0.77<br>(0.33 to 1.21) | 85% | 0.13 (-0.55 to 0.82) |
| ≥ 40<br>(mild) | 2 | 0.60<br>(-0.57 to 1.77) | 83% | 0.13 (-0.06 to 0.32) |

\*The proportional-recovery difference is the between-arm difference in change expressed as a fraction of (66 – baseline FMA-UE).

To examine whether treatment effects scaled with the theoretical room for recovery, we re-expressed the change scores. Each change score was divided by the distance to the scale ceiling (66 − baseline FMA-UE). The pooled between-arm difference in this recovered proportion was 0.11 (95% CI 0.08 to 0.14; P < 0.001). Thus, rTMS-treated patients regained on average an additional 11% of their residual recovery potential. This proportional gain did not vary significantly with baseline severity (meta-regression slope 0.003 per FMA-UE point, P = 0.158). The point estimate was numerically smallest in the severe band (0.03). This again suggests, without establishing, that the most severely impaired patients may benefit least.

### Neurophysiological determinants of treatment response

We addressed two complementary questions. First, does corticospinal tract (CST) integrity, indexed by motor-evoked potential (MEP) status or diffusion-based tract measures, moderate the clinical response? Second, do rTMS-induced changes in corticospinal excitability and conduction parallel functional recovery?

#### CST integrity as a potential moderator of treatment response

Two trials permitted within-trial, biomarker-stratified comparisons (Table 3). Wang L et al., 2022[15] stratified participants by diffusion tensor imaging–derived fractional anisotropy of the CST. The effect was substantially larger in the preserved-CST stratum (SMD 0.72) than in the impaired-CST stratum (SMD 0.02). Qian W et al., 2024[55] stratified participants by MEP status. The MEP-positive stratum showed a large effect (SMD 1.74), whereas the MEP-negative stratum showed essentially none (SMD 0.14). Pooling the two stratified trials (Fig 5), the effect was 1.26 (95% CI 0.27 to 2.25) in preserved-CST/MEP-positive strata. In impaired strata, it was 0.10 (95% CI −0.40 to 0.60). The between-stratum interaction was statistically significant (z = 2.06, P = 0.040).

**Fig 5.**
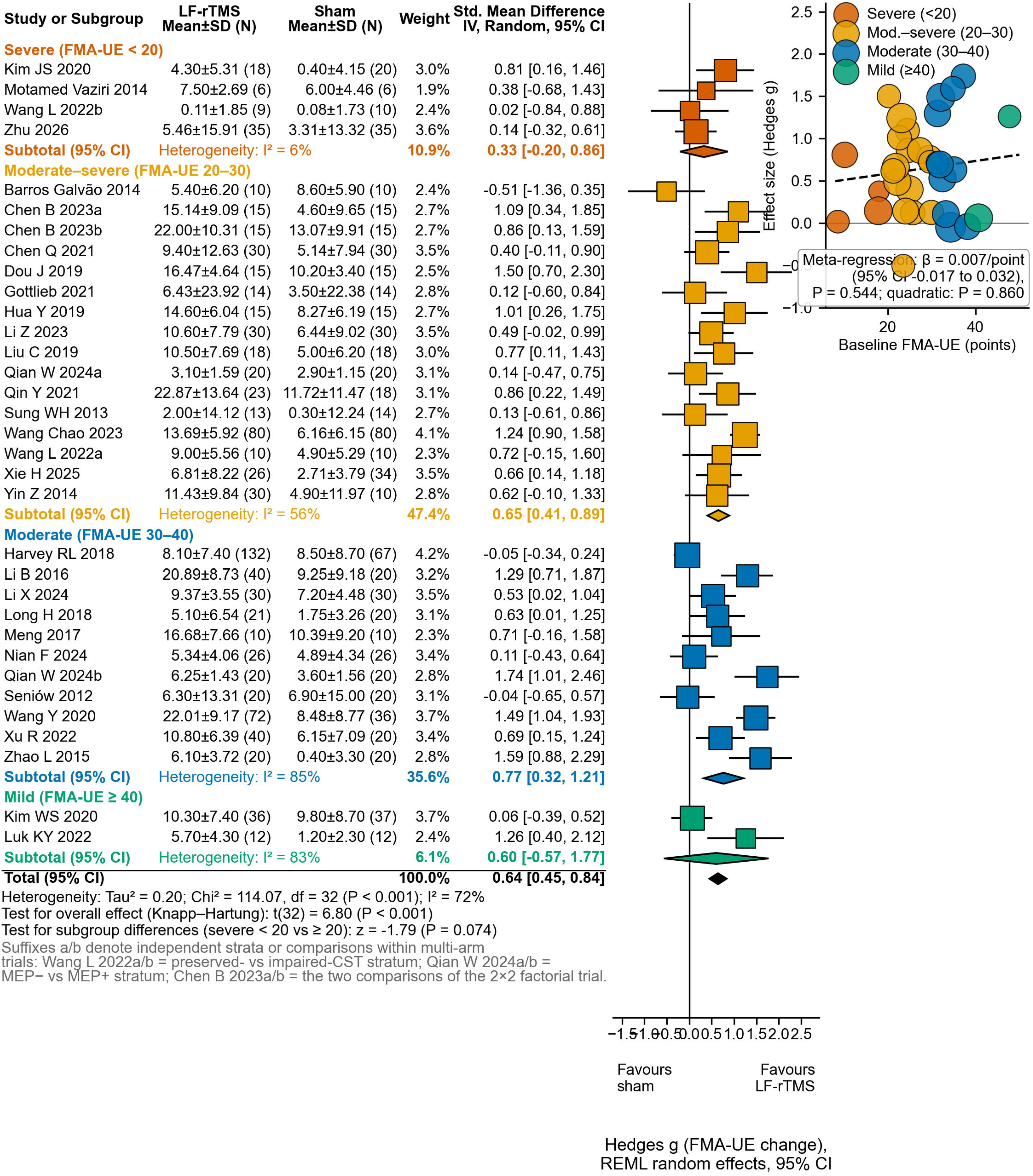
CST integrity as a moderator of treatment response.

**Table 3.** Within-trial, biomarker-stratified effects of contralesional LF-rTMS on FMA-UE change.

| Trial | Biomarker | SMD, preserved stratum | SMD*, impaired stratum | Interaction |
| --- | --- | --- | --- | --- |
| Wang L 2022 | DTI-derived CST fractional anisotropy | 0.72 | 0.02 | $P = 0.278$ (within trial) |
| Qian W 2024 | Ipsilesional MEP status | 1.74 | 0.14 | P = 0.001 (within trial) |
| Pooled (random effects) | CST/MEP integrity | 1.26 (95% CI 0.27 to 2.25) | 0.10 (95% CI -0.40 to 0.60) | z = 2.06, P = 0.040 |

Only two trials contributed to this analysis; the results should be interpreted with caution.
\*SMD values are Hedges g for each stratum-specific comparison; the pooled interaction contrasts the preserved versus impaired strata across the two trials.

Two further observations qualify this pattern. First, [57] enrolled a predominantly severe population (mean baseline FMA-UE ≈ 17.9). It found only a small, non-significant effect (SMD 0.15), consistent with CST-dependent responsiveness. Second, [13] enrolled an extremely severe population (mean baseline FMA-UE 10.2). In that trial, 93% of participants had no recordable ipsilesional MEP. Yet it reported a moderate effect under conservative imputation (SMD 0.81), largely confined to proximal upper-limb movements. This outlier cautions against an absolute CST-dependence rule. At the study level, MEP enrichment was likewise an unreliable proxy. The single trial requiring a recordable ipsilesional MEP at enrollment [59] showed a large effect (SMD 1.24).

However, enrichment status and effect size did not align consistently across the remaining trials. Moderator inferences must therefore rely on within-trial stratification, not on study-level enrichment characteristics.

Taken together, the within-trial stratified evidence supports CST integrity as a moderator of the response to contralesional LF-rTMS. Baseline FMA-UE severity appears to act mainly as a correlated proxy. This conclusion rests on only two biomarker-stratified trials. The certainty of this moderator evidence is therefore very low and should be regarded as hypothesis-generating. The pooled interaction was also only marginally significant (P = 0.040), so this moderator evidence should be regarded as exploratory rather than confirmatory.

#### Electrophysiological changes and their relationship with functional recovery

Neurophysiological outcomes were available for subsets of trials. They were synthesized on the standardized scale because measurement units and protocols varied. For latency-type outcomes measured in milliseconds, mean differences were also computed. Eight comparisons [40,41,46–49,54,59] (546 participants) reported ipsilesional MEP latency.

Eight comparisons [43,46–49,51,59] (468 participants) reported central motor conduction time (CMCT). Six comparisons [13,40,41,43,56] (290 participants) reported MEP amplitude.

Compared with sham, contralesional LF-rTMS significantly shortened ipsilesional MEP latency (SMD 0.95, 95% CI 0.37 to 1.53; P = 0.006; I² = 84%). The mean difference was 1.16 ms (95% CI 0.35 to 1.97). LF-rTMS also shortened CMCT (SMD 1.30, 95% CI 0.58 to 2.03; P = 0.004; I² = 81%). The mean difference was 1.10 ms (95% CI 0.26 to 1.94). MEP amplitude showed a borderline increase that did not reach significance (SMD 0.63, 95% CI −0.01 to 1.26; P = 0.052; I² = 73%). Heterogeneity was substantial for all three measures. This reflects differences in recording muscles, stimulation intensity, and units. Directionally, all three estimates indicate enhanced ipsilesional corticospinal excitability and faster conduction after active stimulation. This pattern is consistent with the intended disinhibition of the ipsilesional hemisphere through suppression of contralesional M1.

The pooled effects on neurophysiological outcomes are presented in Fig 6.

**Fig 6.**
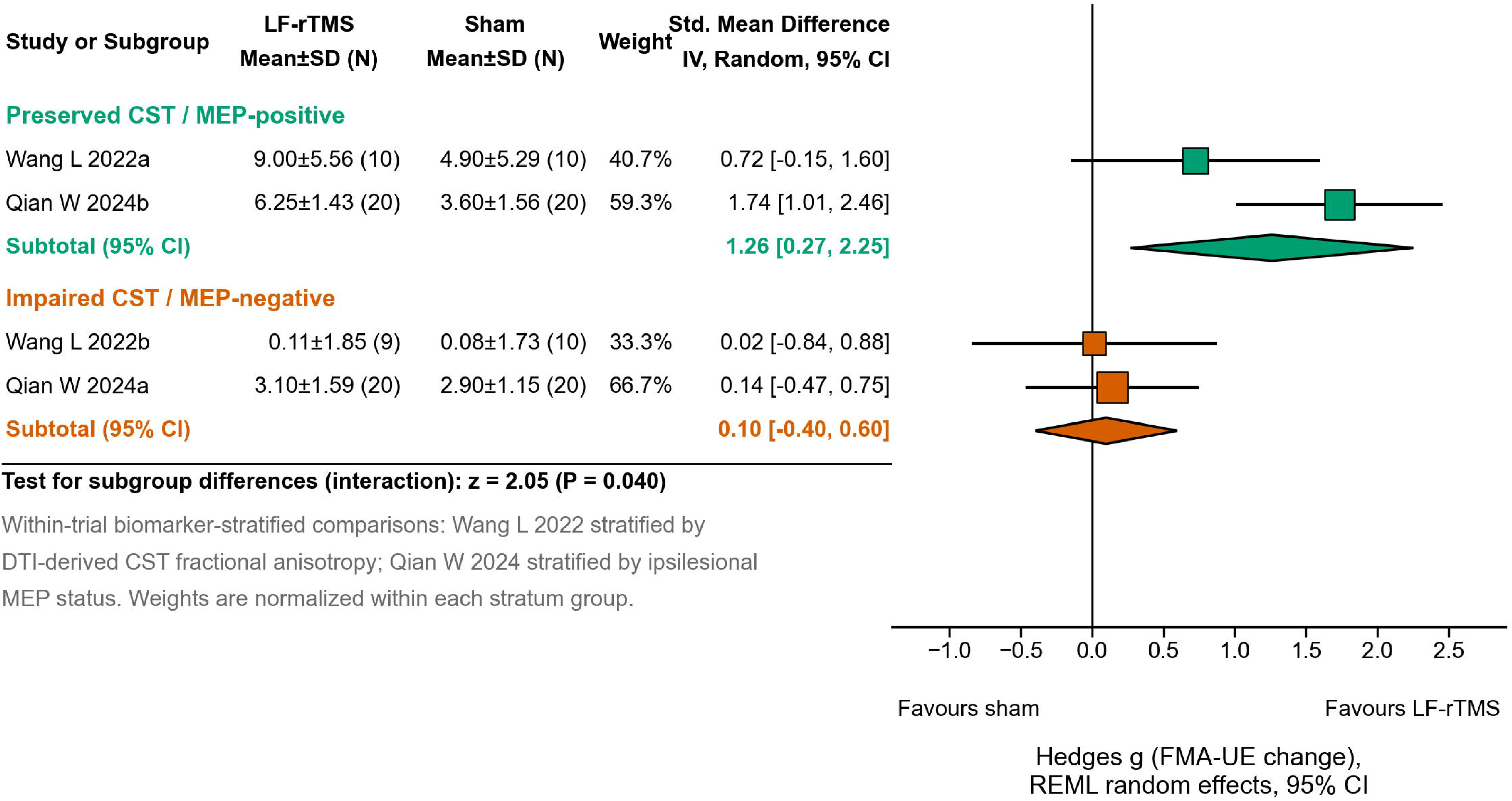
Effects of contralesional LF-rTMS on neurophysiological outcomes.

Exploratory study-level correlations between neurophysiological and functional effect sizes were positive but imprecise for MEP amplitude (Spearman ρ = 0.75, P = 0.084, k = 6). They were essentially null for MEP latency (ρ = −0.36, P = 0.39, k = 8) and CMCT (ρ = −0.08, P = 0.84, k = 8). These analyses use few studies and aggregate-level data. They cannot establish mediation and should be interpreted descriptively. The certainty of evidence was low for MEP latency and CMCT (downgraded for risk of bias and inconsistency). It was very low for MEP amplitude (additionally downgraded for imprecision). A full GRADE evidence profile is provided in Table S3.

#### Summary

Contralesional LF-rTMS produced measurable neurophysiological effects. Ipsilesional MEP latency and CMCT were shorter, and MEP amplitude showed a borderline increase. These directions are congruent with the interhemispheric-inhibition rationale. However, the aggregate-level evidence linking these changes to functional gains remains indirect.

The neurophysiological measures cannot yet be regarded as validated surrogate endpoints. Across the moderator analyses, CST integrity emerged as a more plausible determinant of response than clinical severity itself. The within-trial stratified interaction was significant (P = 0.040). Severity-based modification was null on continuous testing and only weakly suggested by banding. Both conclusions are hypothesis-generating and require confirmation in biomarker-stratified trials. Formal mediation analysis was not feasible with aggregate data from so few studies (k = 6–8). These measures should therefore not currently be used as surrogate endpoints for clinical benefit. Individual-level mediation models within future trials are required to establish surrogacy.

### Sensitivity analyses and robustness of findings

A series of prespecified sensitivity analyses examined the influence of risk of bias, individual studies, and statistical assumptions on the primary result.

#### Influence of risk of bias

Excluding the six studies at high overall risk of bias [37,41,45,49,50,52] left 27 comparisons. The pooled mean difference was 3.75 points (95% CI 2.36 to 5.14; P < 0.001). This is essentially unchanged from the primary estimate of 4.11 points.

Heterogeneity remained substantial (I² = 82%). The primary conclusion is therefore not driven by studies at high risk of bias.

#### Influence of individual studies

In the leave-one-out analysis, pooled mean differences ranged narrowly from 3.69 to 4.29 points. Every iteration remained statistically significant. No single comparison materially influenced the result. Excluding the largest trial ([33] n = 199, terminated early for futility) raised the pooled estimate slightly to 4.29 points (95% CI 3.01 to 5.57).

Excluding Wang Y et al., 2020 yielded 3.69 points (95% CI 2.59 to 4.79). That trial carried a data-quality flag because its methodology section was near-identical to that of Li B et al., 2016.

#### Statistical assumptions

Of the 33 comparisons, 25 required correlation-based imputation of the change-score standard deviation. Two strata of one trial used standard deviations derived from reported 95% confidence intervals [55]. One comparison with an anomalous reported standard deviation (implied pre–post correlation above 0.95) was conservatively imputed in the primary analysis [13]. The choice of imputation correlation had little impact. With r = 0.2, the MD was 3.90 (95% CI 2.66 to 5.13); with r = 0.8, it was 4.34 (95% CI 3.02 to 5.66). This is consistent with the empirical pre–post correlations in the four studies reporting complete SDs. Their median was 0.91 (range 0.74–0.99 across 8 arms), placing r = 0.5 on the conservative side. For [13], using the reported rather than the imputed control-arm SD increased that study’s effect (SMD 1.35 versus 0.81). The pooled estimate was virtually unchanged (SMD 0.66, 95% CI 0.46 to 0.86, versus 0.64, 95% CI 0.45 to 0.84). Alternative between-study variance estimators produced similar pooled effects. DerSimonian–Laird gave an MD of 4.07 (95% CI 2.86 to 5.29) and an SMD of 0.64 (95% CI 0.45 to 0.84). Paule–Mandel gave an SMD of 0.64 (95% CI 0.46 to 0.83). Re-computation under the standardized metric did not alter inference.

#### Summary

Across all sensitivity analyses, the pooled effect remained positive and statistically significant. These analyses included exclusion of high risk-of-bias studies, leave-one-out, and exclusion of the largest trial. They also included exclusion of flagged studies, alternative imputation correlations, and dual handling of the anomalous-SD study.

Finally, they included alternative variance estimators and metric re-computation. Every estimate stayed within approximately 0.5 FMA-UE points of the primary estimate.

Substantial heterogeneity persisted throughout. The direction and approximate magnitude of the primary finding are therefore robust. Its precision and its translation into clinically important benefit remain uncertain, as shown by the wide prediction interval (the sensitivity analysis section). Detailed results are given in Fig S1.

### Adverse events (safety)

Across all included trials, rTMS was generally well tolerated. No severe adverse events were directly attributed to the intervention. The most common adverse effects were transient and mild. They included headache [33,36], scalp discomfort or paresthesia at the stimulation site [41,59], dizziness [58], and transient fatigue. In the largest multicenter trial [33], 26 serious adverse events occurred in 18 of 196 participants. None were deemed related to the study device or intervention, and between-group differences were not significant. No seizures or stroke recurrences were reported in any included study.

Overall, low-frequency rTMS (0.25–1 Hz) delivered within international safety guidelines appears safe and well tolerated after stroke. This is consistent with previous safety assessments [60,61].

### Publication bias

The funnel plot of effect size against standard error was broadly symmetric on visual inspection (Fig 7). Formal tests found no evidence of small-study effects (Egger regression intercept 1.04, P = 0.358; Peters’ test, P = 0.970). The trim-and-fill procedure identified no potentially missing comparisons. The PET-PEESE procedure did not indicate material publication-bias inflation. The PET-adjusted estimate was 0.31 (95% CI −0.35 to 0.98, P = 0.345). The PEESE model was not invoked because the PET intercept was non-significant. Although no statistical signal of publication bias was detected, caution is still warranted. Most trials were small, single-center studies with infrequent prospective registration. Selective reporting at the study level therefore cannot be fully excluded. Moreover, formal tests have limited power when reporting biases operate uniformly across similar-sized studies.

**Fig 7.**
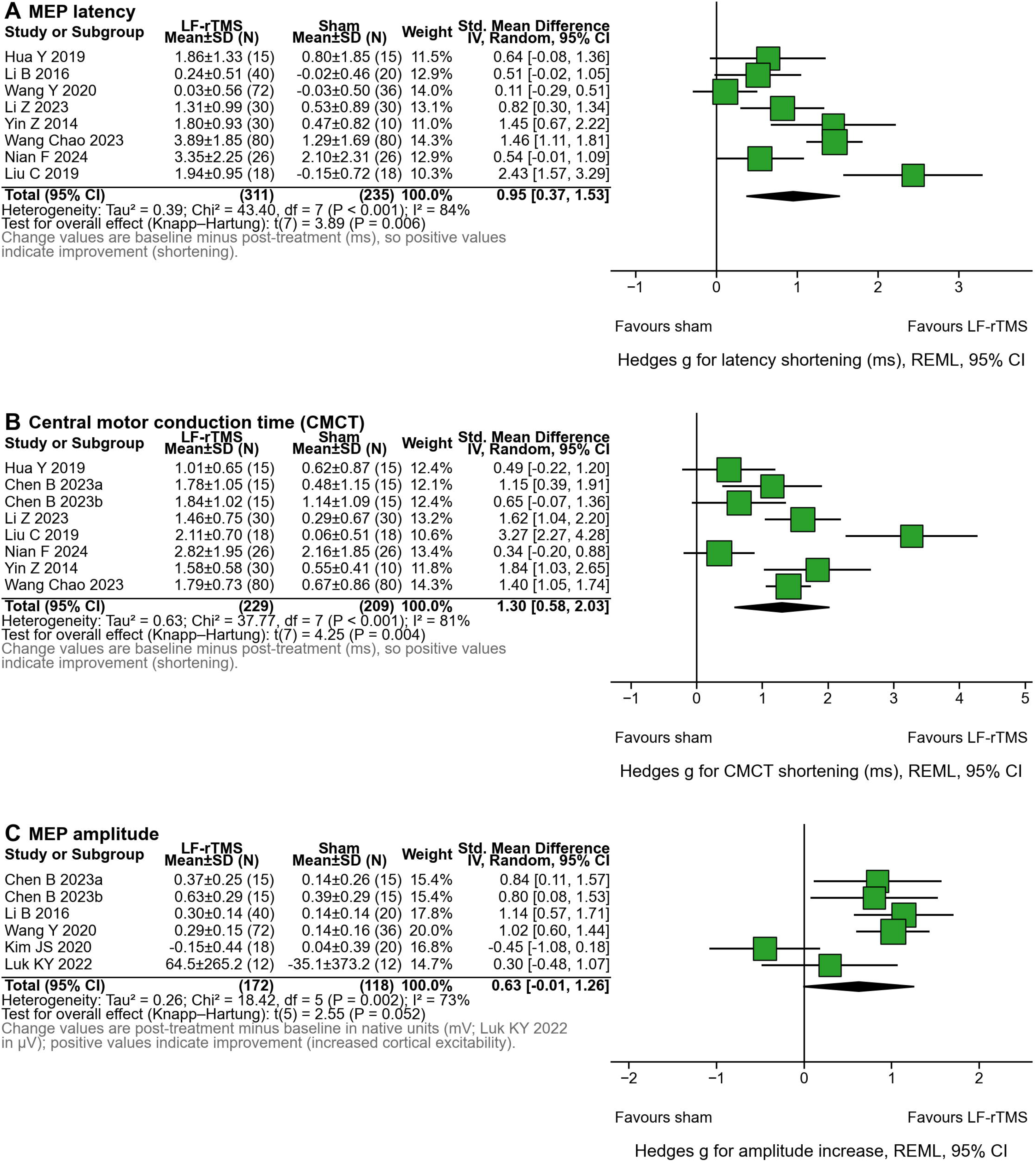
Contour-enhanced funnel plot.

## Discussion

### Integrated interpretation of the main findings

This meta-analysis of 33 sham-controlled comparisons confirms that contralesional LF-rTMS improves upper-limb motor function after stroke. The intervention was superimposed on balanced conventional rehabilitation (MD 4.11 FMA-UE points; SMD 0.64). Three qualifiers define the clinical meaning of this finding. First, the average effect sits just below the conventional minimal clinically important difference of 5–6 points.

The intervention shifts the distribution of recovery in a favorable direction. However, an average patient should not be expected to cross the clinically important threshold from stimulation alone. Second, heterogeneity was substantial and only partly explainable. The 95% prediction interval (−2.32 to 10.54 points) implies genuinely variable effects across settings. The pooled mean should therefore be read as an average over a wide distribution of true effects, not as a uniform benefit. Third, the effect was stable across every sensitivity analysis. The qualitative conclusion, benefit on average, does not depend on any single study, estimator, or imputation choice.

The prespecified effect-modification analyses yielded an internally coherent pattern that challenges two common assumptions in the field. Baseline FMA-UE severity, analyzed continuously, did not modify the treatment effect (linear P = 0.544; quadratic P = 0.860), and the prespecified band contrast between severe and non-severe populations fell short of significance (P = 0.074); that patients with milder impairment simply benefit more is therefore not supported as a general rule. This conclusion converges with the broader literature. Reviews and large multicenter cohorts report no simple “milder-is-better” gradient for LF-rTMS. Where severity effects do appear, they are protocol-dependent and non-linear. This pattern is consistent with our sham-controlled data [62,63]. In contrast, the two trials stratified by corticospinal tract integrity showed a consistent but only marginally significant interaction. The pooled SMD was 1.26 in preserved strata versus 0.10 in impaired strata (P = 0.040). This exploratory pattern raises the possibility that the biological substrate of the stimulation target determines who responds. An intact corticospinal pathway can transmit reorganized descending commands. The clinical severity score itself appears less decisive and acts as an imperfect, correlated proxy. The proportional-recovery analysis points in the same direction. Treated patients regained an additional 11% of their theoretical recovery potential on average, with no significant severity scaling (P = 0.158). This is difficult to reconcile with a pure proportional-recovery account, in which absolute gains should track baseline severity [18,19].

The neurophysiological syntheses showed directionally coherent effects. Ipsilesional MEP latency and CMCT were shorter, and amplitude showed a borderline increase.

These findings support the intended mechanistic action of contralesional inhibition [5,6,8]. Nevertheless, aggregate-level correlations between neurophysiological and functional effects were weak and imprecise, so these measures should currently be viewed as mechanistic markers of target engagement rather than validated surrogate endpoints for functional recovery [16].

### Comparison with previous meta-analyses

Our pooled estimate is directionally consistent with earlier syntheses of rTMS for post-stroke upper-limb recovery [9,10,12]. However, it is more conservative. Several design features account for this difference. We restricted the evidence base to sham-controlled trials with balanced co-interventions. This isolates the net effect of LF-rTMS rather than the combined effect of stimulation plus unequal rehabilitation. We pooled change scores with a conservative imputation correlation (r = 0.5, bracketed by empirical estimates).

We also applied the Knapp–Hartung adjustment, which yields wider but more reliable intervals under substantial heterogeneity. Finally, we included two large pragmatic trials with null results [31,33]. The latter could be synthesized only qualitatively. These trials anchor our estimate closer to the null than meta-analyses dominated by small positive trials.

With respect to effect modification, our findings diverge from a recent meta-analysis that reported stratified effects by baseline severity [11]. That analysis relied on between-study subgrouping without within-trial stratification. In our data, the severity gradient suggested by banding (P = 0.074) was not confirmed by the more powerful continuous meta-regression (P = 0.544). The gradient itself was also confounded by biomarker status. By incorporating the two available biomarker-stratified trials [15,55], this study provides the pooled within-trial evaluation of this question. Specifically, it examines whether corticospinal integrity modifies the response to contralesional LF-rTMS after stroke. The evidence is limited, but the observed interaction is biologically plausible.

CST integrity may explain response variability beyond baseline clinical severity. Finally, network meta-analyses rank multiple stimulation modalities [9]. In contrast, our single-protocol focus avoids transitivity assumptions. It yields a directly interpretable estimate for this specific, widely used protocol.

### Methodological strengths and limitations

This review followed prospective registration and Cochrane and PRISMA standards. Its prespecified moderator framework, conservative estimators, and extensive sensitivity analyses strengthen confidence in the findings. It also provides the first quantitative pooling of biomarker-stratified comparisons for this intervention. Several limitations remain. The moderator analyses used study-level aggregates, so they reflect trial-level associations rather than individual-patient predictors. The severe stratum was thin (k = 4 comparisons), and the CST-integrity interaction rests on only two trials. Heterogeneity was substantial and largely unexplained. In addition, 25 of 33 comparisons required change-score imputation. Finally, only end-of-treatment effects were synthesized. The single available six-month trial reported no maintained advantage [31].

The evidence base was also geographically concentrated: 23 of the 31 included studies were conducted in China (including Taiwan and Hong Kong), with the remaining trials spread across six countries. A country-stratified subgroup analysis, although prespecified, was therefore not informative and is reported as a limitation rather than as a result. This concentration restricts external validity, because rehabilitation routines, stimulation dosing, and reporting practices may differ across regions. It also leaves room for selective reporting that formal statistical tests cannot detect. Replication in non-Asian health-care settings is therefore needed.

The near-absence of long-term follow-up further constrains clinical decision-making. Only one trial reported outcomes beyond the end of treatment, and its six-month result was null [31]. Without extended follow-up, we cannot determine whether the end-of-treatment benefit persists, fades, or accumulates with continued rehabilitation. Future trials should prespecify follow-up assessments of at least three months.

### Clinical and research implications

For practice, contralesional LF-rTMS is a safe adjunct to conventional rehabilitation with a modest average benefit. Clinicians should communicate calibrated expectations, because average gains approximate but do not clearly exceed clinical importance.

Baseline severity alone should not guide patient selection. Where available, corticospinal integrity assessment may help identify responders, pending prospective validation.

However, this exploratory signal should not yet be used to guide treatment selection in routine practice.

Looking forward, biomarker-stratified trials with adequate power for interaction testing are the key next step [15,55]. Individual-participant-data meta-analysis could clarify patient-level predictors, particularly in severe impairment. Standardized neurophysiological endpoints, tested with individual-patient mediation models, would clarify whether neurophysiological changes causally mediate functional gain. Above all, longer follow-up of at least three months is essential: the null six-month result of the E-FIT trial [31] suggests that durability, not immediate efficacy, may decide clinical adoption. Contralesional LF-rTMS thus remains a promising but incompletely targeted intervention, whose average benefit is real yet modest.

## Data Availability

All data produced are available online at https://doi.org/10.5281/zenodo.21889795. All extracted study-level data supporting the meta-analysis results are included within the manuscript and its Supporting information files.

## Author Contributions

Conceptualization: Mianxuan Yu, Man Hao. Data curation: Mianxuan Yu, Jun Lin.

Formal analysis: Mianxuan Yu, Yiming Zeng. Funding acquisition: None.

Investigation: Mianxuan Yu, Jun Lin.

Methodology: Mianxuan Yu, Yiming Zeng, Man Hao.

Project administration: Man Hao.

Resources: Man Hao.

Software: Mianxuan Yu, Yiming Zeng. Supervision: Man Hao, Huanghao Zhou.

Validation: Jun Lin, Yiming Zeng.

Visualization: Mianxuan Yu, Yiming Zeng.

Writing – original draft: Mianxuan Yu.

Writing – review & editing: Mianxuan Yu, Man Hao, Huanghao Zhou, Yiming Zeng, Jun Lin.

## Acknowledgments

The authors have no acknowledgments to declare.

## Supporting information

Table S1. Search strategy.

Table S2. Detailed RoB 2 assessments.

Table S3. GRADE summary of findings.

Table S4. Sensitivity analysis results.

Table S5. PRISMA 2020 checklist.

Fig S1. Sensitivity analyses.

---

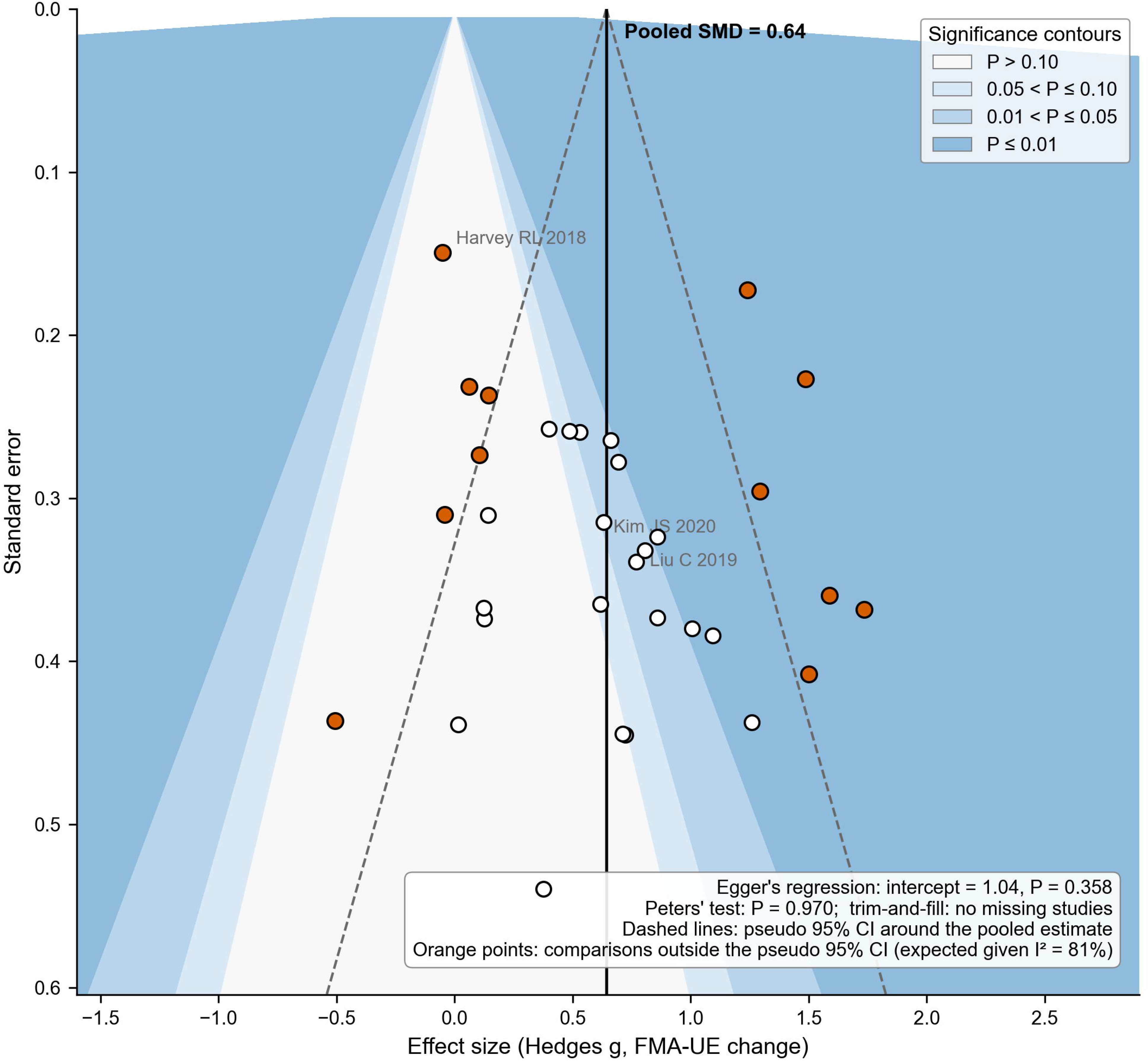

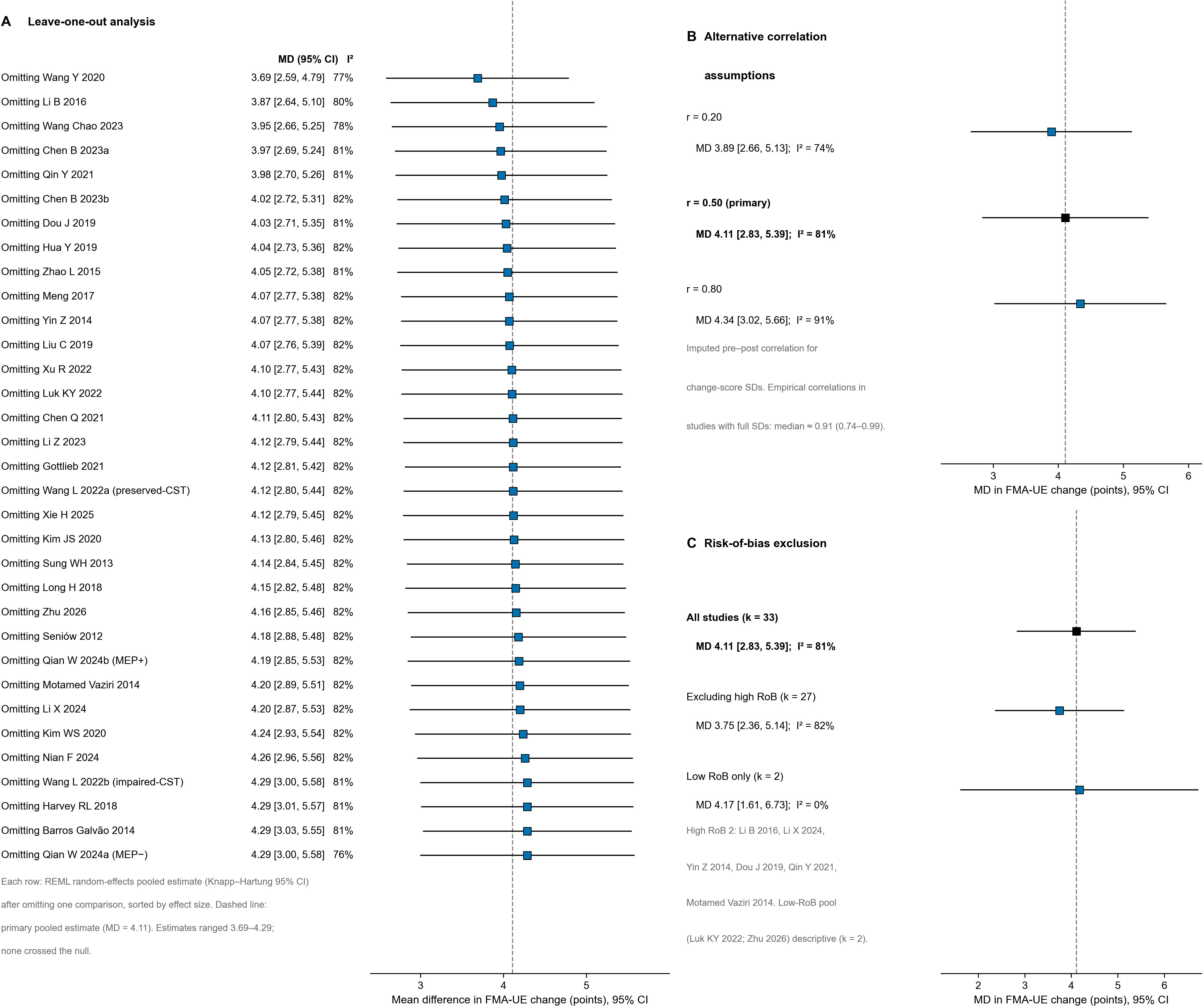

