## Supplementary material for "Clinical and neurophysiological determinants of response to contralesional low-frequency repetitive transcranial magnetic stimulation after stroke: A systematic review and meta-analysis": Table S1

**Table S1. Search strategy**

| Database | Search strategy |
| --- | --- |
| PubMed | ("Stroke"[MeSH] OR stroke[tiab] OR poststroke[tiab] OR "cerebrovascular accident"[tiab]) AND ("Transcranial Magnetic Stimulation"[MeSH] OR rTMS[tiab] OR "repetitive transcranial magnetic stimulation"[tiab]) AND ( "Upper Extremity"[MeSH] OR upper limb[tiab] OR arm[tiab] OR hand[tiab] OR "motor recovery"[tiab] OR "motor function"[tiab])AND (randomized controlled trial[pt] OR random*[tiab]) |
| Embase | ('stroke'/exp OR stroke:ti,ab OR poststroke:ti,ab OR 'cerebrovascular accident':ti,ab)AND('transcranial magnetic stimulation'/exp OR rTMS:ti,ab OR 'repetitive transcranial magnetic stimulation':ti,ab)AND('upper limb'/exp OR 'upper extremity':ti,ab OR arm:ti,ab OR hand:ti,ab OR 'motor recovery':ti,ab OR 'motor function':ti,ab)AND ('randomized controlled trial'/exp OR random*:ti,ab) |
| Web of Science | TS=(stroke OR poststroke OR "cerebrovascular accident") AND TS=("transcranial magnetic stimulation" OR rTMS OR "repetitive transcranial magnetic stimulation") AND TS=("upper limb" OR "upper extremity" OR arm OR hand OR "motor recovery" OR "motor function") AND TS=(random* OR randomized OR RCT) |
| Cochrane CENTRAL | ( MeSH descriptor:[Stroke] explode all trees ) AND ( MeSH descriptor:[Transcranial Magnetic Stimulation] explode all trees )  AND ( MeSH descriptor:[Upper Extremity] explode all trees ) |
| CNKI | (主题:卒中+脑卒中+中风+脑梗死+脑梗塞+脑出血+偏瘫)  AND (主题:重复经颅磁+rTMS+低频重复经颅磁刺激+低频rTMS) AND (主题:上肢+上肢功能+上肢运动功能+FMA+Fugl-Meyer) |
| Wanfang | 主题=(脑卒中 OR 卒中 OR 脑血管意外 OR 脑梗死 OR 脑缺血 OR 脑出血 OR 缺血性卒中 OR 出血性卒中) AND (重复经颅磁刺激 OR 经颅磁刺激 OR rTMS OR TMS OR 重复性经颅磁刺激) AND (上肢 OR 上肢功能 OR 上肢运动功能 OR 手功能 OR 肢体功能 OR 运动功能 OR 运动恢复 OR 运动障碍) AND (随机 OR 随机对照 OR 随机分组 OR 对照试验 OR 临床试验) |
| VIP | 篇关摘=(脑卒中 OR 卒中 OR 脑血管意外 OR 脑梗死 OR 脑缺血 OR 脑出血 OR 缺血性脑卒中 OR 出血性脑卒中) AND (重复经颅磁刺激 OR 重复性经颅磁刺激 OR 经颅磁刺激 OR rTMS OR TMS) AND (上肢 OR 上肢功能 OR 上肢运动功能 OR 手功能 OR 运动功能 OR 运动恢复 OR 肢体功能) AND (随机 OR 随机对照 OR 随机分组 OR 对照试验 OR 临床试验) |
