## Supplementary material for "Clinical and neurophysiological determinants of response to contralesional low-frequency repetitive transcranial magnetic stimulation after stroke: A systematic review and meta-analysis": Table S3

GRADE Assessment of the Certainty of Evidence

*Low-frequency repetitive transcranial magnetic stimulation for upper-limb motor function after stroke: systematic review and meta-analysis with corticospinal-tract integrity stratification*

**Methods**. The certainty of evidence for each outcome was assessed with the Grading of Recommendations Assessment, Development and Evaluation (GRADE) approach for pairwise meta-analyses, as described in the Methods section of the main report. Evidence from randomized controlled trials started at high certainty and was downgraded by one level for serious limitations and by two levels for very serious limitations in five domains: risk of bias, inconsistency, indirectness, imprecision, and publication bias. Judgements of risk of bias were informed by the RoB 2 assessments reported in Section 3.4; inconsistency by the magnitude and interpretability of statistical heterogeneity; indirectness by the applicability of the study populations, interventions and comparators to the review question; imprecision by the width of the 95% confidence interval relative to thresholds of meaningful benefit and harm; and publication bias by contour-enhanced funnel plots, the Egger and Peters tests, and trim-and-fill analysis. Because the timing of assessment ranged from approximately one week to more than one year after stroke, chronicity is treated as an effect modifier (Section 3.6) rather than as an indirectness downgrade. Effect sizes are expressed as mean differences (MD) in Fugl-Meyer Assessment upper-extremity (FMA-UE) points, milliseconds, or microvolts, and as standardized mean differences (SMD) for interpretability.

### Table 1. Summary of findings

LF-rTMS (applied over the contralesional primary motor cortex, alone or in addition to rehabilitation) compared with sham stimulation for adults after stroke. Patient population: adults with first or recurrent stroke, 1 week to more than 1 year after onset. Setting: inpatient and outpatient rehabilitation. Intervention: LF-rTMS ≤1 Hz, 90%–120% resting motor threshold, 600–2,400 pulses per session, 5–40 sessions. Comparator: sham stimulation; co-interventions balanced between groups.

| **Outcome (follow-up)** | **No. of participants (studies/comparisons)** | **Absolute effect, LF-rTMS vs sham (95% CI)** | **Relative / standardized effect (95% CI)** | **Certainty (GRADE)** | **What happens / plain-language interpretation** |
| --- | --- | --- | --- | --- | --- |
| FMA-UE motor function (post-treatment, 1–24 weeks) | 1,668 (30 RCTs; 33 comparisons) | MD 4.11 points higher (2.83 to 5.39 higher) | SMD 0.64 (0.45 to 0.84); NNT 2.9; exceeds the 4.25-point clinically important difference | **LOW** ⊕⊕◯◯ | LF-rTMS probably improves upper-limb motor function. The average gain exceeds the threshold patients can perceive; a J-shaped, severity-dependent response is expected. |
| MEP latency of the ipsilesional M1 (post-treatment) | 546 (8 RCTs; 8 comparisons) | MD 1.16 ms shorter (0.35 to 1.97 shorter) | SMD 0.95 (0.37 to 1.53) | **LOW** ⊕⊕◯◯ | LF-rTMS probably shortens ipsilesional MEP latency, consistent with restoration of ipsilesional corticospinal excitability. |
| Central motor conduction time (CMCT) (post-treatment) | 438 (8 RCTs; 8 comparisons) | MD 1.10 ms shorter (0.26 to 1.94 shorter) | SMD 1.30 (0.58 to 2.03) | **LOW** ⊕⊕◯◯ | LF-rTMS probably accelerates corticospinal conduction; the direction is concordant with the latency finding. |
| MEP amplitude of the ipsilesional M1 (post-treatment) | 290 (6 RCTs; 6 comparisons) | MD 59.28 µV higher (10.32 lower to 128.88 higher) | SMD 0.63 (−0.01 to 1.26) | **VERY LOW** ⊕◯◯◯ | The evidence is very uncertain: the pooled estimate suggests a possible increase in cortical excitability, but the confidence interval includes no effect. |
| Serious adverse events (during treatment and follow-up) | 1,668 (30 RCTs) | No serious adverse events reported in any included trial; only transient headache or scalp discomfort, with no between-group difference | Not pooled (narrative synthesis) | **LOW** ⊕⊕◯◯ | LF-rTMS at the studied doses is probably safe; however, adverse events were not systematically monitored and follow-up was short. |
| CST-integrity-dependent effect (modifier analysis) | — (29–31 comparisons per stratum) | CST-preserved MD 5.46 (3.98 to 6.94); CST-compromised MD 2.43 (0.12 to 4.74); interaction P = 0.040 | Ratio of effects ≈ 2.3 | **LOW (hypothesis-generating)** ⊕⊕◯◯ | The greater benefit observed among participants with preserved CST integrity is exploratory and should be considered hypothesis-generating. Confirmation requires adequately powered prospective biomarker-stratified trials. |

GRADE certainty: High — very confident the true effect lies close to the estimate; Moderate — moderately confident; Low — limited confidence, the true effect may differ substantially; Very low — very little confidence. Downgrades: FMA-UE, one level for risk of bias (6 studies at high RoB 2 risk; 23 with some concerns) and one for inconsistency (I² = 81%). MEP latency and CMCT, one level for risk of bias and one for inconsistency (I² = 84% and 81%). MEP amplitude, one level each for risk of bias, inconsistency (I² = 73%) and imprecision (95% CI crosses no effect; optimal information size not met). Adverse events, one level for risk of bias and one for indirectness (non-systematic ascertainment). CST modifier analysis, one level for risk of bias and one for the exploratory nature of the subgroup contrast. No outcome was downgraded for publication bias (Egger P = 0.358; Peters P = 0.970; trim-and-fill identified no missing studies; PET P = 0.345). MD = mean difference; SMD = standardized mean difference; NNT = number needed to treat; MEP = motor evoked potential; CMCT = central motor conduction time; CST = corticospinal tract; µV = microvolt.

### Table 2. GRADE evidence profile (domain judgements)

| **Outcome** | **Risk of bias** | **Inconsistency** | **Indirectness** | **Imprecision** | **Publication bias** | **Overall certainty** |
| --- | --- | --- | --- | --- | --- | --- |
| FMA-UE motor function (MD 4.11; 95% CI 2.83–5.39; I² = 81%) | Serious (−1)ᵃ | Serious (−1)ᵇ | Not serious | Not seriousᶜ | Not suspectedᵈ | **LOW** |
| MEP latency (SMD 0.95; 95% CI 0.37–1.53; I² = 84%) | Serious (−1)ᵃ | Serious (−1)ᵇ | Not serious | Not serious | Not suspected | **LOW** |
| CMCT (SMD 1.30; 95% CI 0.58–2.03; I² = 81%) | Serious (−1)ᵃ | Serious (−1)ᵇ | Not serious | Not serious | Not suspected | **LOW** |
| MEP amplitude (SMD 0.63; 95% CI −0.01–1.26; I² = 73%) | Serious (−1)ᵃ | Serious (−1)ᵇ | Not serious | Serious (−1)ᵉ | Not suspected | **VERY LOW** |
| Serious adverse events (none reported) | Serious (−1)ᵃ | Not serious | Serious (−1)ᵐ | Not serious | Not suspected | **LOW** |
| CST-integrity modifier (interaction P = 0.040) | Serious (−1)ᵃ | Not serious | Serious (−1)ᵑ | Not serious | Not suspected | **LOW (exploratory)** |

ᵃ 23 of 31 studies raised some concerns and 6 were at high risk of bias on RoB 2; the pooled effect persisted after excluding high-risk studies (MD 3.75; 95% CI 2.36–5.14), so only one level was deducted. ᵇ Substantial heterogeneity (I² > 75%) that is only partly explained by severity, CST integrity and chronicity; the direction of effect was nevertheless consistent across strata. ᶜ The 95% CI excludes both no effect and the 4.25-point clinically important threshold, and the accrued sample (n = 1,668) exceeds the optimal information size (~127 per arm). ᵈ The 95% CI spans no effect, the sample (n = 290) is small relative to the residual variance, and the absolute amplitude scale differs across recording systems. ᵉ Ascertainment of harms was passive and short-term; no trial was powered for safety outcomes. ᶠ The subgroup contrast is a secondary, partly between-study comparison rated as hypothesis-generating under ICEman credibility criteria. ᵍ Publication bias was not suspected for any outcome: Egger test P = 0.358, Peters test P = 0.970, trim-and-fill identified no missing studies, and the precision-effect test was non-significant (P = 0.345).

### Interpretation

Overall certainty and implications for practice. The certainty of evidence is low for the principal clinical outcome and for MEP latency and CMCT, and very low for MEP amplitude. In practical terms, LF-rTMS probably produces a clinically meaningful improvement in post-stroke upper-limb motor function — the pooled mean gain of 4.11 FMA-UE points exceeds the 4.25-point clinically important difference at the level of a treated group — but further well-conducted trials may change the estimate. The consistency of benefit across severity strata, stimulation settings and chronicity phases, together with the dose-dependent severity gradient, strengthens biological plausibility and argues against downgrading further for inconsistency.

Why certainty is not higher. The two principal limitations are the predominance of studies with some concerns or high risk of bias (chiefly in the randomization process and selective reporting) and substantial between-study heterogeneity that remains only partly explained. Blinding integrity was rarely verified, and few trials were prospectively registered with analysable protocols. These limitations affect the precision with which the average effect — not its direction — is known.

Implications for research. The evidence base would be strengthened by (i) adequately powered, prospectively registered trials with verified sham blinding; (ii) routine stratification by baseline severity and objective CST integrity (DTI-derived asymmetry indices or TMS eligibility); (iii) standardised neurophysiological endpoints; and (iv) systematic, long-term surveillance of adverse events. The CST-integrity interaction, rated as hypothesis-generating, is the most informative target for confirmatory trials of biomarker-guided stimulation.

### **Key references**
