## Supplementary material for "Clinical and neurophysiological determinants of response to contralesional low-frequency repetitive transcranial magnetic stimulation after stroke: A systematic review and meta-analysis": Table S4

**Table S4. Sensitivity analysis results for the primary outcome (Fugl-Meyer Assessment upper-extremity change score)**

| **Sensitivity analysis** | **k** | **MD, points (95% CI)** | **SMD, Hedges g (95% CI)** | **I², %** | **P value** |
| --- | --- | --- | --- | --- | --- |
| *Primary analysis and model specification* | | | | | |
| Primary analysis (REML estimator, Knapp–Hartung adjustment) | 33 | 4.11 (2.83 to 5.39) | 0.64 (0.45 to 0.84) | 81 | < 0.001 |
| Alternative τ² estimator: DerSimonian–Laird | 33 | 4.07 (2.86 to 5.29) | 0.64 (0.45 to 0.84) | 81 | < 0.001 |
| Alternative τ² estimator: Paule–Mandel | 33 | 4.10 (2.82 to 5.37) | 0.64 (0.46 to 0.83) | 81 | < 0.001 |
| Common-effect (fixed-effect) model | 33 | 2.58 (2.15 to 3.02) | 0.60 (0.50 to 0.70) | 81 | < 0.001 |
| *Change-score SD imputation assumptions* | | | | | |
| Pre–post correlation r = 0.20 (primary: r = 0.50) | 33 | 3.89 (2.66 to 5.13) | 0.53 (0.37 to 0.69) | 74 | < 0.001 |
| Pre–post correlation r = 0.80 (primary: r = 0.50) | 33 | 4.34 (3.02 to 5.66) | 0.94 (0.66 to 1.21) | 91 | < 0.001 |
| Kim JS 2020 with reported (non-imputed) change SDsᵃ | 33 | 4.11 (2.84 to 5.38) | 0.66 (0.46 to 0.86) | 81 | < 0.001 |
| *Influence of risk of bias and individual studies* | | | | | |
| Excluding 6 studies at high risk of bias (RoB 2)ᵇ | 27 | 3.75 (2.36 to 5.14) | 0.60 (0.38 to 0.82) | 82 | < 0.001 |
| Excluding the largest trial (Harvey RL 2018, n = 199)ᶜ | 32 | 4.29 (3.01 to 5.57) | 0.67 (0.48 to 0.86) | 81 | < 0.001 |
| Excluding Wang Y 2020 (data-quality flag)ᵈ | 32 | 3.69 (2.59 to 4.79) | 0.61 (0.42 to 0.80) | 77 | < 0.001 |
| Leave-one-out analysis (33 iterations)ᵉ | 32 | range 3.69 to 4.29 | — | 76–82 | all < 0.001 |

Values are pooled estimates from random-effects meta-analysis (REML) with Knapp–Hartung 95% confidence intervals unless otherwise indicated; DerSimonian–Laird, Paule–Mandel and common-effect rows use normal-theory intervals. k = number of comparisons; MD = mean difference in FMA-UE change (points); SMD = standardized mean difference (Hedges g). ᵃ Reported change SDs implied a pre–post correlation > 0.95; the primary analysis used conservative imputation (r = 0.50). ᵇ Li B 2016, Li X 2024, Yin Z 2014, Dou J 2019, Qin Y 2021, Motamed Vaziri 2014. ᶜ Terminated early for futility. ᵈ Change-score dispersion flagged as implausibly small relative to baseline and post-treatment SDs. ᵉ Each iteration pools the remaining 32 comparisons.
